# How demographic factors matter for antimicrobial resistance – quantification of the patterns and impact of variation in prevalence of resistance by age and sex

**DOI:** 10.1101/2023.09.24.23296060

**Authors:** Naomi R Waterlow, Ben S Cooper, Julie V Robotham, Gwenan M Knight

## Abstract

**Background:** Antibiotic usage, contact with high transmission healthcare settings as well as changes in immune system function all vary by a patient’s age and sex. Yet, most analyses of antimicrobial resistance (AMR) ignore demographic indicators and provide only country level resistance prevalence values.

In this work we use routine surveillance data on serious infections in Europe to characterise the importance of age and sex on incidence and resistance prevalence patterns for 33 different bacteria and antibiotic combinations. We fit Bayesian multilevel regression models to quantify these effects and provide estimates of country-, bacteria- and drug-family effect variation.

**Results:** At the European level, we find distinct patterns in resistance prevalence by age that have previously not been explored in detail. Trends often vary more within an antibiotic family than within a bacterium: clear resistance increases by age for methicillin resistant *S. aureus* (MRSA) contrast with a peak in resistance to several antibiotics at ∼30 years of age for *P. aeruginosa.* This diverges from the known, clear exponential increase in infection incidence rates by age, which are higher for males except for *E. coli* at ages 15-40.

At the country-level, the patterns are highly context specific with national and subnational differences accounting for a large amount of resistance variation (∼38%) and a range of associations between age and resistance prevalence. We explore our results in greater depths for two of the most clinically important bacteria–antibiotic combinations. For MRSA, age trends were mostly positive, with 72% of countries seeing an increased resistance between males aged 1 and 100 and more resistance in males. This compares to age trends for aminopenicillin resistance in *E. coli* which were mostly negative (males: 93% of countries see decreased resistance between ages 1 and 100) with more resistance in females. A change in resistance prevalence between ages 1 and 100 ranged up to ∼0.46 (95% CI 0.37 – 0.51, males) for MRSA but varied between 0.16 (95% CI 0.23-0.3, females) to -0.27 (95%CI -0.4 - - 0.15, males) across individual countries for aminopenicillin resistance in *E. coli*.

**Conclusion:** Prevalence of resistance in infection varies substantially by the age and sex of the individual revealing gaps in our understanding of AMR epidemiology. These context-specific patterns should now be exploited to improve intervention targeting as well as our understanding of AMR dynamics.

## Introduction

Antimicrobial resistance (AMR) is a global public health priority (1). Understanding how it will be affected by the dramatic demographic shifts that are underway worldwide is a key knowledge gap. The WHO has estimated that one in five people in the world will be aged 60 years or older by 2050 (2). Incidence of bacterial infections is known to increase by age (3) and vary by sex, though the exact trends are rarely quantified. The higher burden of infection in older age groups, results in higher antibiotic exposure and higher contact with healthcare settings which are known hotspots of resistant bacteria transmission. However, there is not a simplistic increase in resistance in all pathogens by age. Determining how the above interact to drive the dynamics of drug resistant infections (DRI) is a vital part of understanding how best to tackle AMR.

Age- and sex-disaggregated data are collected by most routine AMR surveillance schemes. The WHO Global Antimicrobial Resistance and Use Surveillance System (GLASS) requests age- and sex-stratifications from reporting countries (4). However, this data is not openly available at low age-band segregation (i.e. more than 4 broad categories) with sex – for example not from the WHO GLASS dashboard (5) nor the ECDC Surveillance ATLAS dashboard (6) nor the US Centres for Disease Control and Prevention (7). The recent WHO reports also have no presented analysis of how resistance prevalence varies by these demographic factors (8,8). The dramatic, often exponential, increase in infection incidence with older age has been reported in several places (9–12) as well as the differences by sex (13). However, how this burden is split into resistant or susceptible infection by patient age and sex is relatively rarely reported. Multiple attempts to predict AMR burden are hampered by a basic lack of surveillance data and yet factors such as sex and age are variables that are nearly always available for analysis.

The importance of age being linked to variation in AMR has been graphically explored before for Europe (14) and more comprehensively in single setting studies (e.g. (15–17)). Complex statistical analysis based on the Global Burden of Disease methods have produced age- and sex-specific estimates of mortality rates by European country attributable to all AMR (18). However, to our knowledge there has been no comprehensive analysis of the relationship between age and AMR in infection between bacteria across multiple countries. This is despite the wide awareness of age-specific effects for infection that have only been emphasised by the COVID-19 pandemic (19).

Despite awareness of the importance of sex for many risk factors for infectious diseases (e.g. HIV) and bacterial infections such as urinary tract infections, how prevalence of drug-resistant infection (DRI) varies between the sexes (and genders) is vastly underexplored in the literature (20). This is despite many studies of single bacteria or syndromes finding a difference in resistance prevalence in infection between the sexes (21–26). In 2018, the WHO called for countries to take the first step to better considering “gender and equity” in National Action Plans for AMR (27), which have historically lacked such considerations (e.g. in Southeast Asia (28)).

Prevalence of resistance in infection is known to vary between countries (5,8) and sub-nationally, by factors such as governance and deprivation level (29–31). This may be linked in part to differences in healthcare structures and antibiotic usage (32,33). Other national level healthcare structures and cultural differences are likely to have wider impacts on AMR patterns by age and sex. For example, variation in birth rates by age between countries (34), as well as type of birth (vaginal vs caesarean)(35) will impact the type of antibiotic as well as healthcare exposures in women. Determining how these cultural factors interplay with biological factors as we age and across sexes is key to understanding the complexity of AMR interventions.

Here, we use large, routinely collected data on bloodstream infection to explore generalised trends in antibiotic resistance prevalence in infection by age and sex in Europe. It is vital for antibiotic prescribing to account for local trends in resistance, as well as how they vary by age and sex. We do not wish to oversimplify any trends in AMR by age or sex – risk factors, previous prescribing as well as contact with high-risk transmission settings such as hospital or long-term care facilities will all influence individual level risk of AMR infection. However, our ecological analysis deepens our understanding of AMR epidemiology in Europe and suggests testable hypotheses about the underlying cultural and biological factors driving differences in resistance by age and sex.

## Methods

Our analysis was in three stages – firstly we explored the trends in resistance prevalence by age and sex across Europe, and secondly estimated and quantified the incidence of infection for each of the bacterial species by age and sex. Thirdly, we quantified the proportion of those infections that were due to resistant bacteria for different bacteria-antibiotic combinations by age and sex, country and sub-national indicator (laboratory). All code is available in online repository (36). Patient level data is available upon request from the European Antimicrobial Resistance Surveillance Network (EARS-Net) from the Surveillance System (TESSy) (37).

We show the overall results for trends and incidences across all bacteria-antibiotic combinations, highlighting bacteria-antibiotic combinations of interest, then demonstrate specific age-associated results using two of the clinically most important bacteria-antibiotic combinations: MRSA and *E.coli* – aminopenicillins (amoxicillin and ampicillin).

### Data

We analysed individual European Antimicrobial Resistance Surveillance Network (EARS-Net) patient level data from the Surveillance System (TESSy), provided by Austria, Belgium, Bulgaria, Cyprus, Czechia, Germany, Denmark, Estonia, Greece, Spain, Finland, France, Croatia, Hungary, Ireland, Iceland, Italy, Lithuania, Luxembourg, Latvia, Malta, Netherlands, Norway, Poland, Portugal, Romania, Sweden, Slovenia, Slovakia and the United Kingdom and released by ECDC (37). Countries were anonymised using a random 3-letter code, which is used throughout the paper. This surveillance network collects routine clinical antimicrobial susceptibility testing (AST) results, alongside some patient data, including sex and age, from EU and EEA countries (we use the term European throughout). The general quality and comparability of the data is evaluated through a standard annual external exercise (38) with the AST results taken from shared protocols (39,40). The data consists of AST for the first blood and/or cerebrospinal fluid isolate (< 0.7% of this dataset) of every patient with an invasive infection associated with one of the pathogens under surveillance. Levels of coverage are discussed and explored in the calculation of incidence (see below and Appendix S1, section 3). In our main analysis we exclude individuals aged 0, due to their stark difference in immune dynamics and contact patterns, but run a sensitivity analysis including them.

Individual patient data from EARS-Net was extracted with information on the age and sex of the patient, resistance presence, laboratory code, year of sample and reporting country. For incidence calculations we included all isolates with recorded age and sex values, for those aged one or older (89% of the original 3,549,617 isolates). For resistance prevalence calculations, we used the susceptibility test result data for 2015-2019 in those aged one or older, with data on age and sex. This resulted in a dataset of susceptibility results for 17 antibiotics, 8 bacteria in 29 countries (Appendix S1, section 2). The number of isolates provided from each of the 29 countries varied between 15,298 and 619,648 (Appendix S1, section 2). An analysis on missing data both in terms of (a) distribution of age and sex within those not tested for resistance and (b) resistance prevalence in those without age and sex information is in (Appendix S1, section 2).

We used the United Nations subregion definitions, except for Cyprus, which was grouped with Southern Europe (instead of being the only Western Asia country). Some susceptibility data grouped results for multiple antibiotics together: “aminopenicillins” are ampicillin or amoxicillin, “3G cephalosporins” are cefotaxime, ceftriaxone or ceftazidime, “fluoroquinolones” are ciprofloxacin, levofloxacin or ofloxacin, “aminoglycosides” are gentamicin or tobramycin, “macrolides” are azithromycin, clarithromycin or erythromycin, “penicillins” are penicillin or oxacillin, “carbapenems” are imipenem/meropenem. See ECDC report Table 1 for further bacterial-antibiotic specific pairing (41). Where we had multiple susceptibility results for individual antibiotic within a drug family (beta-lactam) we grouped antibiotics by AWaRE classifications (42). We follow the ECDC analysis and assume “sex” rather than “gender” was recorded in the data.

**Table 1:**
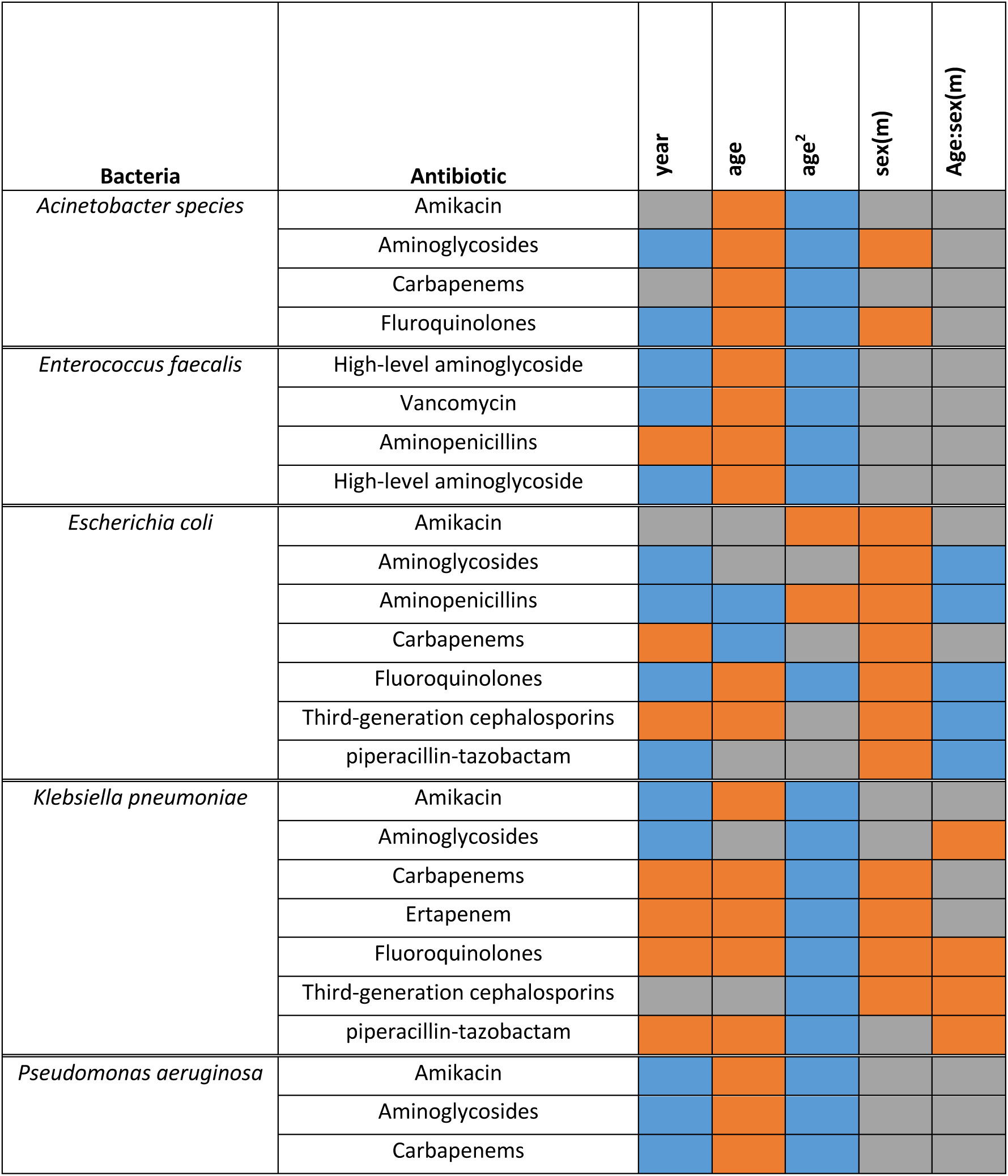
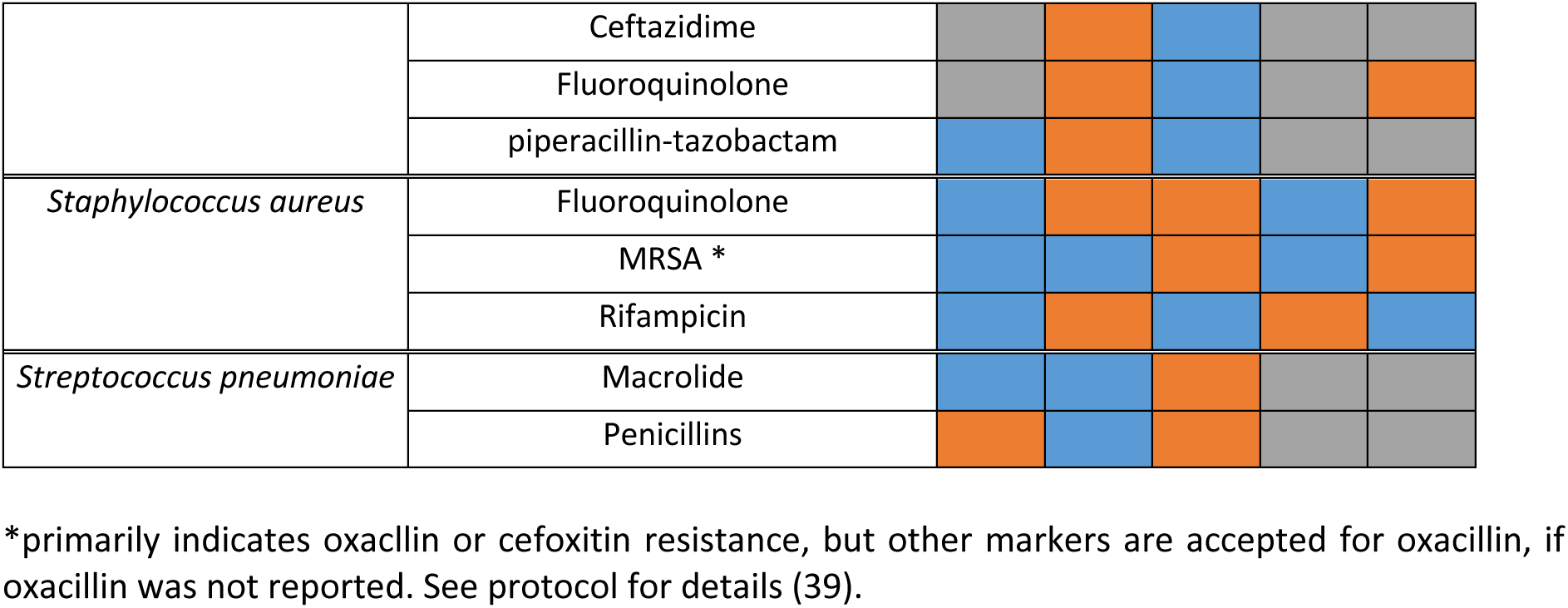
Heatmap of the values of the fixed effect parameters for each bacteria-antibiotic model. Orange indicates a positive effect and blue indicates a negative effect (in both cases, where the 95% credible intervals of the posterior parameter estimate do not cross 0). Grey indicates no effect (i.e. posterior CI’s cross 0. An equivalent table with the parameter values can be found in the supplement (Appendix S2, section 4). (m) indicates that the parameter is the coefficient for males.

### (1) Prevalence of resistance in infection by age and sex

Using the cleaned data we explored variation in patterns in aggregated sex- and age-based resistance prevalence in infection at the European and subregional levels.

### (2) Incidence of infection by age

Following the methods of Cassini et al (43) (Appendix S1, section 3&4), it was assumed that all eligible invasive isolates are reported by the participating laboratories. The estimated coverage of these laboratories was then used as an inflation factor to calculate the number of BSIs. Data for country coverage was taken from previous EARS-Net reports and the Cassini et al estimates for 2015, 2018, 2019 and 2020. The incidence of infection in each of these years was calculated by dividing the number of isolates from patients in each 5yr age and sex band by the corresponding population sizes from the World Bank DataBank (44), up to a pooling of all those aged 80 or older.

### (3) Trend analysis for resistance proportion by age

Multilevel regression models were fitted to the ECDC data to understand the impact of including age and sex in models of resistance prevalence. We used a Bayesian framework using the *R* package *brms* (45) and ran models using the No U-turn Sampling separately for each bacteria-antibiotic combination, using data from 2015-2019. Individual level data was aggregated to group level by country, laboratory code, sex, age and year of sample and standardised as appropriate (Appendix S1, section 5). Models were considered converged if the Rhat was <1.1, a sufficient Effective Sample Size for each parameter was reached and we checked for divergent transitions (Appendix S1 section 5). We initially ran 3000 iterations and extended this to 5000 for those models that had not reached convergence at this point. Country and laboratory code were included as substantial variation was observed between them (Appendix S1, section 5). Only the sexes “male” and “female” were included in the analysis and records missing age or sex were dropped.

Thus, for each bacteria-antibiotic combination, our data consisted of multiple groupings of individual samples of a bacterium tested for resistance to that antibiotic. Each grouping *i* had a unique combination of country (c), laboratory code (l), sex, age and year of sample and hence a linked number of samples (*n*) and proportion resistant (*p*).

For each bacteria-antibiotic combination we ran a multi-level logistic regression model to predict the probability of an isolate being resistant to the antibiotic, assuming a Binomial distribution over the number of samples in each grouping. Our model included both age and sex terms (Equations 1:2).

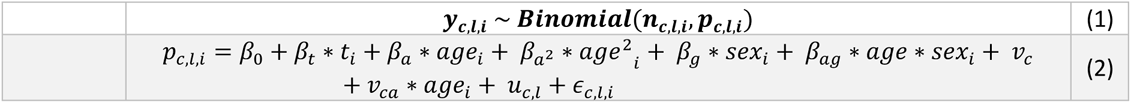

Where *y* is the resistance variable, taking a value of 0 or 1, *n* is the number of samples and *p* the probability of the sample being found to be resistant (NAs were excluded, Appendix S1, section 5). The subscripts *c*, *l* and *i* denote country, laboratory code and grouping level. *β*_0_ is the overall intercept, *β_t_* is slope coefficient for time and *t* is year. *ε_c,l,"_* is the residual error, *u_c,l_* is the level-2 random error on laboratory code and *v_c_* is the level-3 random error on country. *β_a_* is the age effect coefficient, *β_a^2^_* is the age squared effect coefficient and **v_ca_** is the country level age effect coefficient. *β_g_* is the sex effect coefficient and *β_ag_* is the sex and age interaction coefficient. All random errors are assumed to be normally distributed, and we assume flat priors on all covariates in the main analysis, but run a sensitivity analysis with weakly informative regularising priors. We did not use weakly informative priors for the main analysis, as we had no prior information on what the parameter values should be.

To determine an overall impact of age for each bacteria-antibiotic combination and country, we calculated the difference in proportion resistant between individuals aged 1 and those aged 100, using the posterior predictions from the model fit. We did this across all posterior samples, from which we calculated the median and 95% quantiles.

### Sensitivity analysis

We explored further data disaggregation of incidence by patient location when the sample was taken (inpatient vs. outpatient and the hospital unit or ward type e.g. haematology or emergency department) For incidence analysis, we explored varying the inflation factor for the incidence of infection to check robustness of age and sex patterns.

For the modelling analysis, we explored including samples from individuals aged, including regularising priors and using a model selection-based philosophy. These sensitivity analyses were run for MRSA.

### Ethics

This analysis of patterns of resistance in samples taken as part of routine surveillance was approved by the London School of Hygiene and Tropical Medicine Ethics board (ref 28157).

## Results

### European level

At the European level, there were clear non-linear differences in the prevalence of resistance in infection by age and sex for different bacteria-antibiotic combinations (Figure 1). These patterns were robust across subregions of Europe (Appendix S2, section 1). However, prevalence of resistance was generally higher in Southern and Eastern Europe, with stronger age-related trends (e.g. for methicillin resistance in *S. aureus* and across *Acinetobacter spp.*). The age-associated patterns varied more within drug-families than within certain bacteria (patterns within each colour are more different than within each row of Figure 1). For example, patterns of resistance across drug families were highly similar for some bacteria such as Acinetobacter species (Figure 1) whilst there was substantial variation within resistance proportions by age for fluroquinolones (blue data, Figure 1). Sex has little impact on many of the age-related trends except for Acinetobacter species at younger ages, and *E. coli* and *Klebsiella* at higher ages (Figure 1).

**Figure 1:**
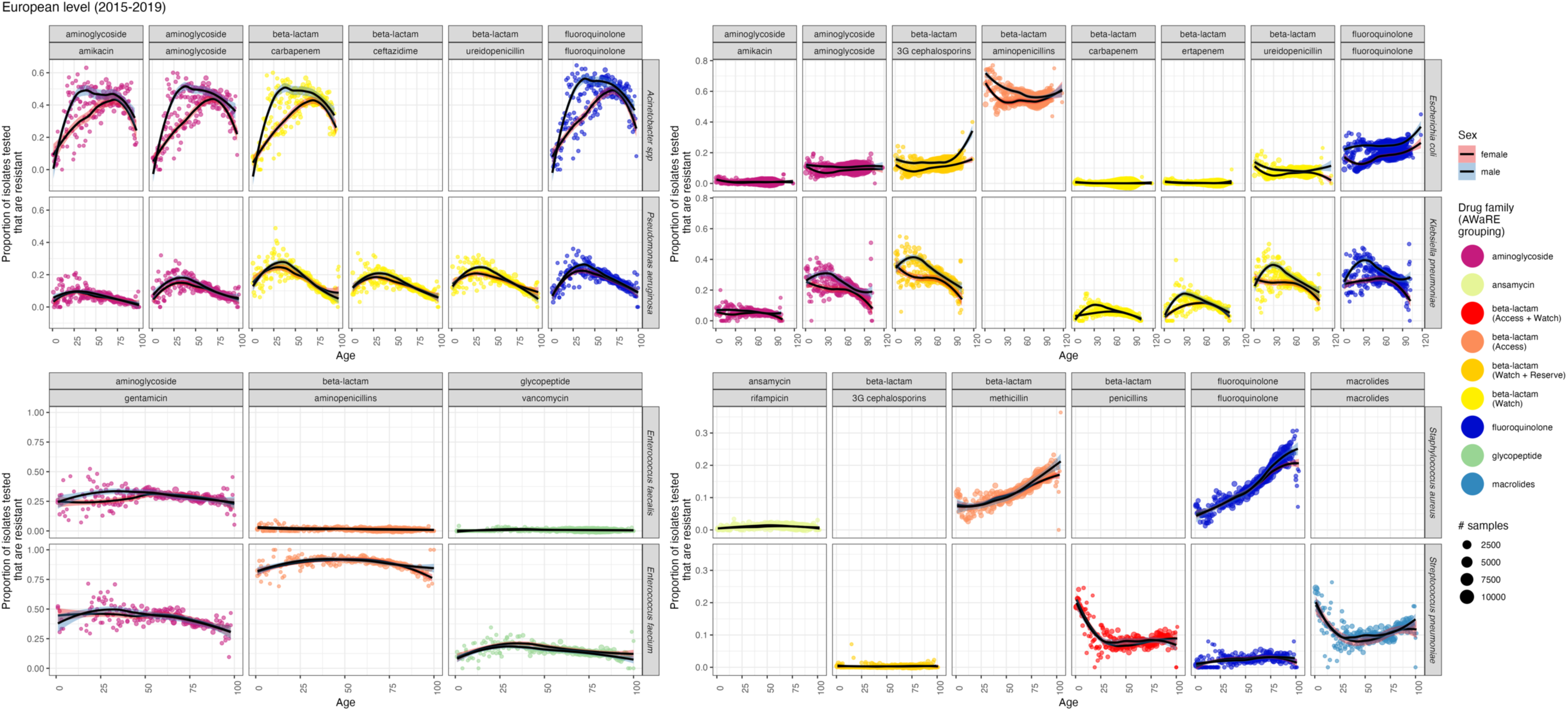
Trends in resistance prevalence vary by antibiotic, bacteria and demographic factors across Europe. The proportion of isolates tested (y axis) that are resistant to each antibiotic (column) within drug families and AWaRE groupings (colour) for each bacteria (row) is shown for all European data over 2015-2019 by age (x axis). Data is shown as points with number of samples indicated by size of point. Shaded areas are 95% confidence intervals around the LOESS fit line by sex (linetype and shade). AWaRE groupings were used here to better distinguish clinically important subsets within the beta-lactam family.

### Incidence

As expected, across Europe, bloodstream infection incidence substantially increased with age with clear differences between the sexes (Figure 2). Men had a higher incidence of infections from approximately age 35 onwards, except for *E. coli* between ages 15 and 40 where women had a higher incidence. These patterns were robust at the country level and over time (Appendix S2, section 2). Differences in infection incidence between the pathogens reflect the overall burden in infection, with ranking incidence rates being (from highest): *E. coli, S. aureus, K. pneumoniae, E faecium, P. aeruginosa, E. faecalis, S. pneumonia, Acinetobacter species*.

**Figure 2:**
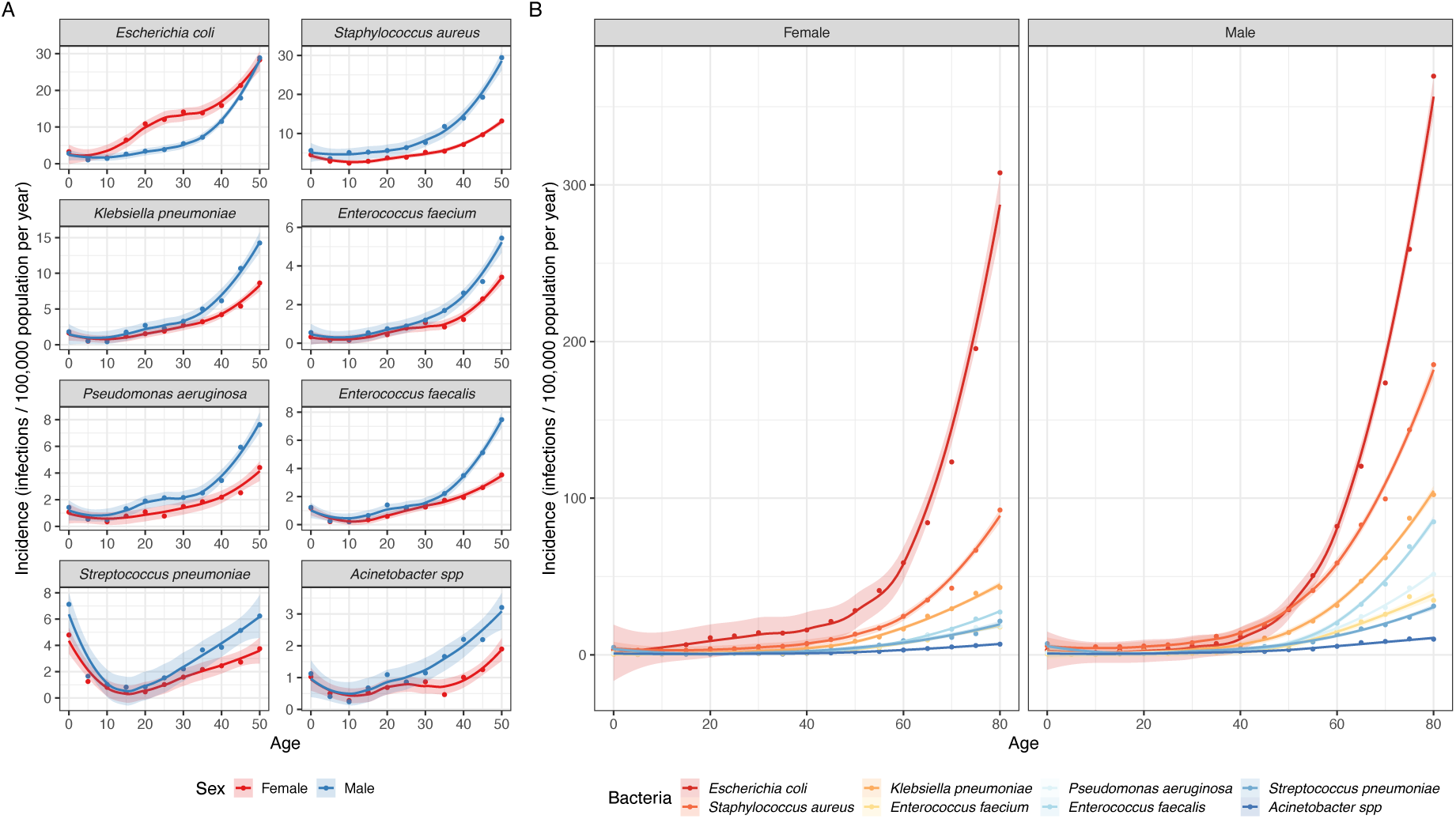
Incidence of bloodstream infections per 100,000 population in 2019 across European countries for 8 bacterial pathogens split by (A) sex and bacteria for the first 50yrs of life and (B) sex (panel) and bacteria (colour) lifelong. Shaded areas are 95% confidence intervals using a LOESS fit. Infections in individuals younger than 0 are excluded, and those aged 80 and older are pooled into the 80yr data point.

The combination of these age-related trends in number of infections (Figure 2) with those in proportion resistant (Figure 1) lead to exponential increases in the number of resistant infections with age, as would be expected (Appendix S2, section 3) for all bacteria-antibiotic combinations.

### Resistance prevalence

Our logistic model converged for 33 bacteria-antibiotic combinations (92%) (Appendix S2, section 4) and supported clear benefits of including age in predictions of resistance in infection, with substantial effects for at least one of age, age^2^ or the interaction between age and sex (Table 1). Sex had less of a clear importance for many bacteria-antibiotic combinations, with at least one of the sex intercept or interaction terms being substantial for 19 of the 33 bacteria-antibiotic combinations. Full results for all bacteria-antibiotic combinations can be found in Appendix S3.

### Model analysis: examples

For MRSA, most countries (e.g. for males, 72%, 21/29 countries) had a positive trend with age (Figure 3), with a higher proportion of samples being resistant at age 100 than age 1 (Figure 3). The magnitude of this difference varied but reached a maximum difference in proportion of ∼0.46 (95% quantile 0.37 – 0.51, country PUB) between males aged 1 and 100. Country level effects (panels, Figure 3) as well as laboratory (subnational) effects (lines, Figure 3) were highly important in capturing proportion resistant by age. There were also significant sex effects in both the intercept and age-slope terms for MRSA, which resulted in an increased age-impact in males (Figure 3).

**Figure 3:**
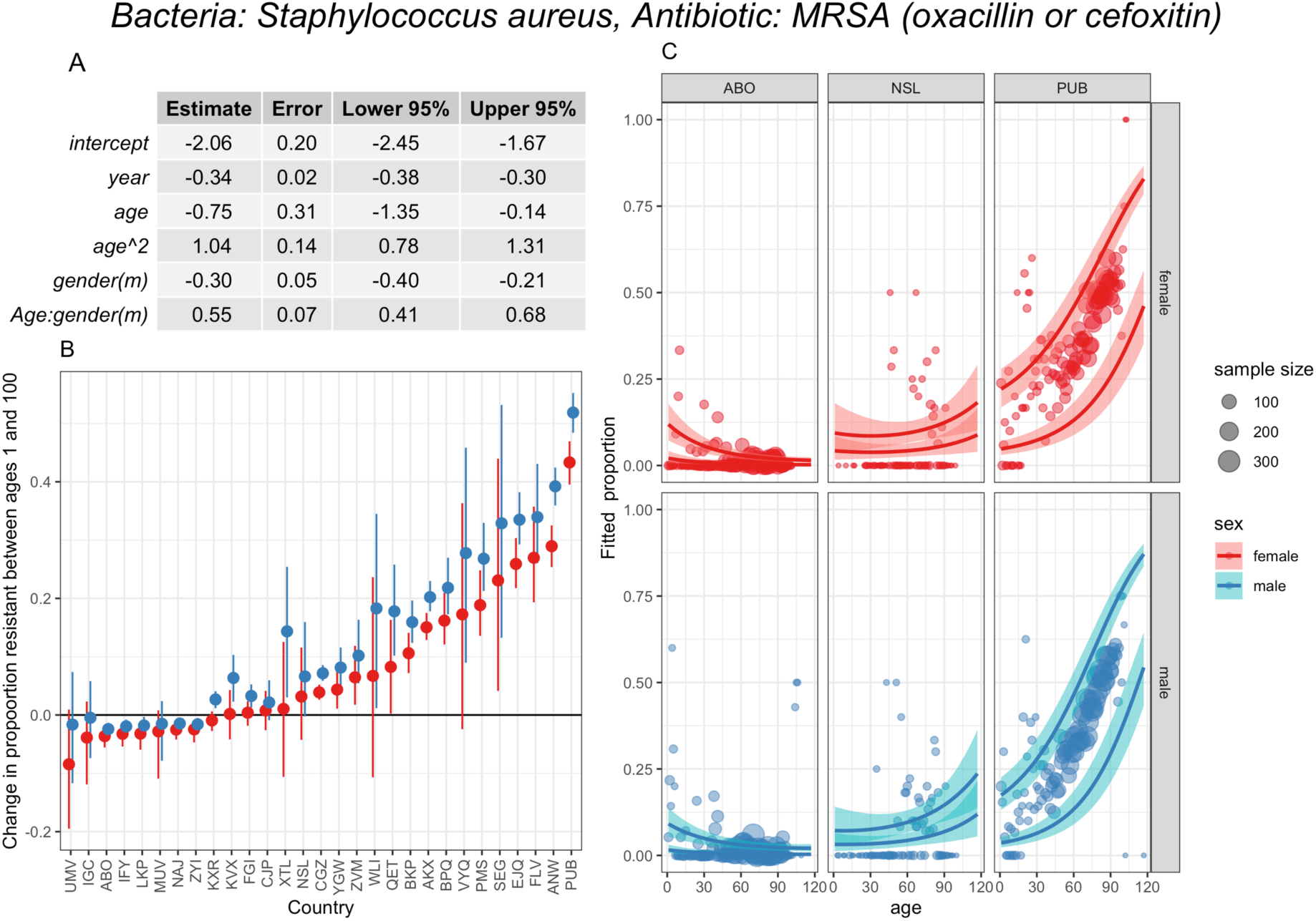
Model analysis for MRSA. A) Model parameters. B) Change in proportion resistant between ages 1 and 100 for each country and sex, with the point indicating the median and the error bars the 95% quantiles. C) Data (points) and model predictions (lines) with 95% CIs (ribbons) for the two sexes (rows) for the most extreme (left&right columns) and the middle country (middle column) estimated age slope. Each country has two lines, depicting the predictions for the most extreme laboratories in the country. Data sample size is grouped across years and laboratories. MRSA primarily indicates oxacllin or cefoxitin resistance, but other markers are accepted for oxacillin, if oxacillin was not reported. See protocol for details (39). Country labels are a random anonymised three letter code used for this analysis only.

For aminopenicillin (amoxicillin and ampicillin) resistance in *E.coli* we also see age effects across countries, however for this bacteria-antibiotic combination the age trend is mostly negative (males, 93%, 27/29 countries), with a lower proportion resistant with age. The magnitude of the age effect varied from 0.16 (95% quantiles 0.12 – 0.20, country BZT, female) to -0.27 (95% quantiles -0.4 - - 0.15, country ABO, male). As with MRSA, substantial country and laboratory variation is observed (Figure 4). There is also a significant sex effect (Figure 4): we found, across ages, a higher proportion resistant in females.

**Figure 4:**
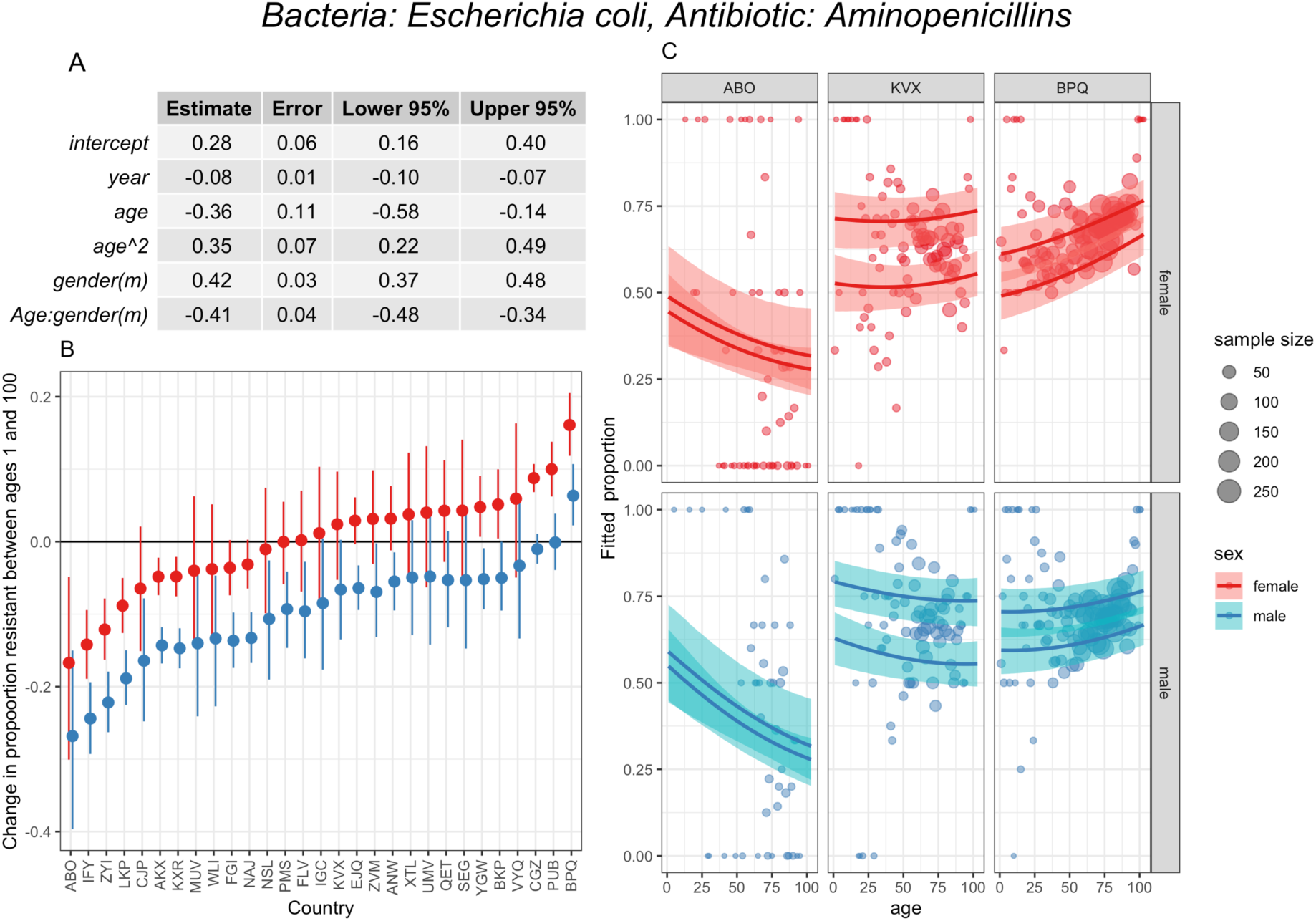
Model analysis for E.coli, aminopenicillin resistance. A) Model parameters. B) Change in proportion resistant between ages 1 and 100 for each country and sex, with the point indicating the median and the error bars the 95% quantiles. C) Data (points) and model predictions (lines) with 95% CIs (ribbons) for the two sexes (rows) for the most extreme (left&right columns) and the middle country (middle column) estimated age slope. Each country has two lines, depicting the predictions for the most extreme laboratories in the country. Data sample size is grouped across years and laboratories. Country labels are random anonymised three letter code used for this analysis only.

### Model analysis: general results

Whilst the two bacteria-antibiotic samples chosen above show substantial trends, there are many bacteria-antibiotic combinations where no age or sex trend is seen, or where there is little cross-country variation (completely overlapping confidence intervals when looking at the age 1 – 100 change) (Appendix S2, section 5). Additional bacteria-antibiotic combinations where a > 5% change in resistance proportion for multiple countries was seen include third-generation cephalosporin resistance in *E coli* and *K. pneumoniae,* fluroquinolone resistance in *E coli, K. pneumoniae,* and *S. aureus*, aminoglycoside resistance in *E. coli* and *K. pneumoniae,* and carbapenem resistance in *P. aeruginosa.* The latter is relatively stable at ∼-10% across countries, whilst others have large variability. There is also substantial variation between different bacteria-antibiotic combinations within countries.

Looking across bacteria-antibiotic combinations, age-related trends were seen across drugs for *Acinetobacter* species (positive, both sexes), *E.coli* (positive, female) and *Pseudomonas aeruginosa* (negative, both sexes) (Figure 5A), although for these the majority were not significant at the 95% level, and have relatively small impacts. No clear age-related trends were linked to Gram stain, nor within antibiotic (or antibiotic families) were seen in our modelling results (Figure 5B), further emphasising the trends seen at the European level.

**Figure 5:**
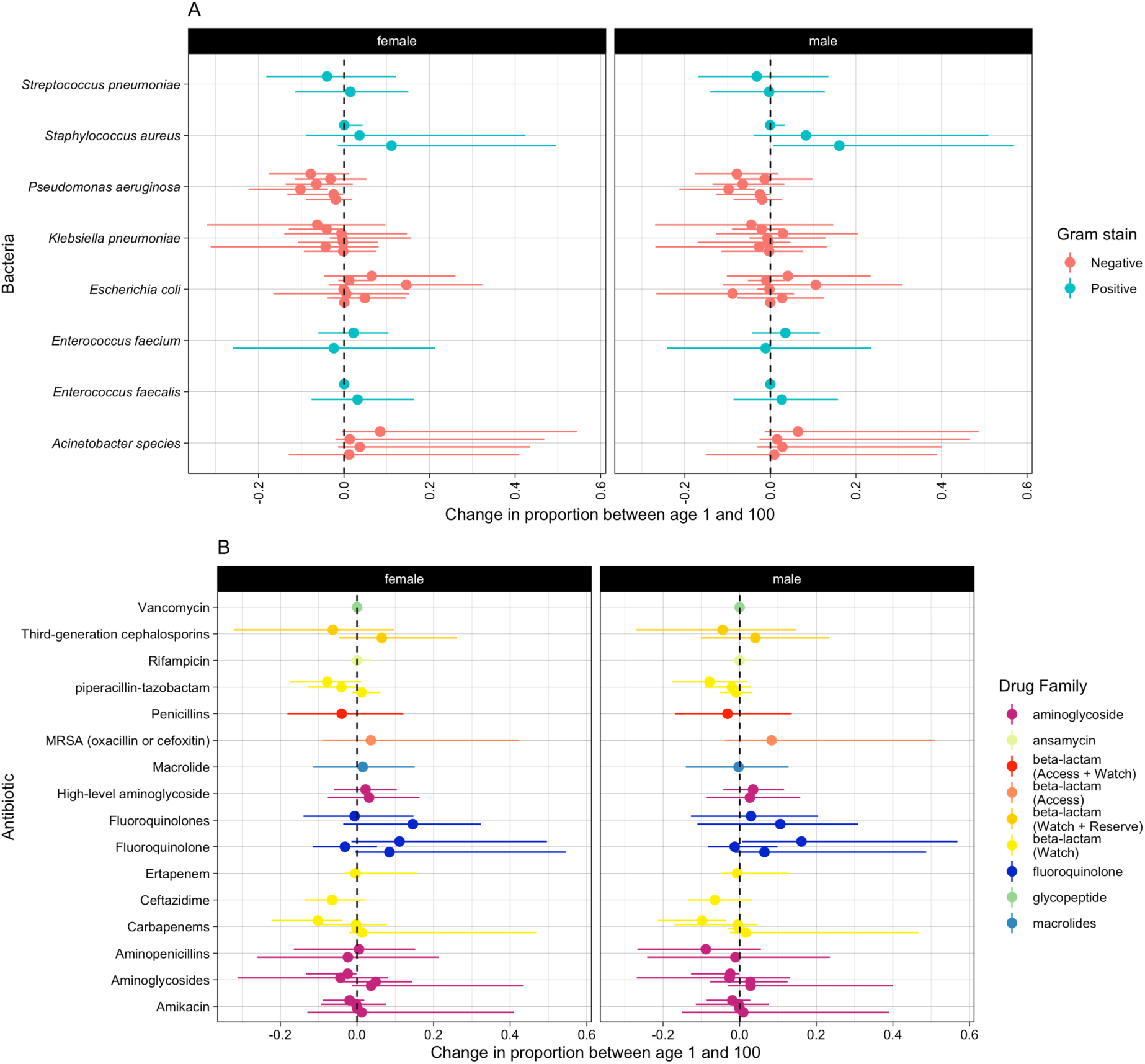
Change in proportion of samples resistant between age 1 and 100, A) by bacteria and B) by drug and drug family. The dashed line indicates 0.

### Sensitivity analysis

Analysis of infection incidence by patient location when the sample was taken revealed large differences between countries across Europe likely linked to differences in healthcare systems and reporting protocols (Appendix S2, sections 6&7). Hence, we could not here explore resistance prevalence further by these differences. Age and sex patterns in incidence were robust to using the minimum surveillance coverage values (Appendix S2, section 8).

Modelling sensitivity analyses showed little effect of including samples from individuals aged 0 or from including regularising priors (Appendix S2, sections 9 & 10). Our model selection sensitivity analysis showed that even with the different philosophical approach, the model used in the main analysis was the preferred model (Appendix S2, section 11).

## Discussion

Our comprehensive quantification of the strong ecological relationships between age and sex with resistance prevalence in bloodstream infection (BSI) across Europe highlights novel relationships not previously studied and suggests that such differences should be considered in AMR research and policy. Whilst there are limited previous studies looking at age and sex in the context of AMR, these do not provide their estimates of the relationships as output (18) or are limited to explorations in specific settings or bacteria (15,24,46–48). The only previous report considering age differences in detail focuses only on 6 bacteria-antibiotic combinations in 5 year age bands, and shows similar trends as we do (14) but did not quantify such differences nor explore them at the national, or sub-national, level.

We find no universal trends, with variation in age and sex patterns across specific bacteria-antibiotic combinations and across national and sub-national contexts, suggesting that cultural factors dominate biological ones. The main strengths of our analyses lie in the detailed BSI data used, as we were able to use one-year age bands, where trends may have been obscured in previous studies by high-level age groupings (e.g. (6,15,18,49)).

Focusing on two critical pathogens for bloodstream infections (*E. coli* (20.5%) and *S. aureus* (20.7%)) (50) and associated important resistances in Europe (41) and globally (51), we show substantial subnational and national variation, whilst demonstrating that there are age- and sex-related trends for specific bacteria-antibiotic combinations. Transmission of MRSA often occurs in healthcare settings (52) and increased contact with such settings with age could explain our observed often positive trend in resistance proportions by age. Whilst for aminopenicillin resistance in *E. coli* the contrasting dominant negative trend in resistance with age could be explained if, with age, more infections were endogenous and community-onset (see below).

Whilst resistance prevalence in many gram-negative bacteria often peaked at younger ages (as has been predicted for multidrug-resistant *M. tuberculosis* carriage (53)), we found little commonality in patterns between bacteria or within drug families. This suggests that the link between demographics and AMR is likely to be less driven by biological factors, but instead is driven more by cultural factors, as alternatively we might expect similar patterns of resistance within drug families across different bacteria. Contrasting this with the biological factors that can drive increased infection risk by age suggests that there is vital information in comparing and contrasting AMR prevalence in infection spatially by age and sex to improve intervention design and antimicrobial usage. For example, understanding trends in resistance by age could lead to improved understanding of the importance of antimicrobial use variation between ages and countries (54), healthcare contact and infection prevention control practices, and even microbiological sampling that could inform both data analysis for burden and evolution understanding as well as transmission intervention potential.

Our results could inform policy and practice in healthcare settings in a variety of ways. Firstly, understanding differences in age- and sex-related risks of infection with resistant bacteria could lead to more targeted empiric prescribing, tailored to the individual and setting, as has previously been suggested (15,17,55). This may be particularly important in older adults, that often experience more severe consequences of bacterial infection (56). When considering empiric prescribing guidelines, it is vital to not just consider the impact of prescribing the most appropriate drug, but also the impact of delaying prescription until more information is available, which may have catastrophic consequences in severe infections <u>(Girard and Ely 2007)</u>. Secondly, understanding the importance of demographic factors on AMR will support the collection and smart use of further data in this area. This is especially relevant due to the high levels of variation across settings that we identify, underpinning the need for local level infection data collection and policies. Whilst demographic data is often encouraged to be reported by countries, this is often not included in analyses, and it’s use is confounded by differences in the data sources and sampling practices (57). These differences, and the variation we found, highlight the need for reducing the reliance on estimates of AMR based on either single settings within a country or national averages, as done with large global estimation studies (49), as averaging across data collected from different study sites can reduce the accuracy and poorly reflect heterogeneity.

This is not only true for understanding AMR, but also BSI risks – sex-differences in incidence by age could give clues to targeting this large contributor to mortality that are not commonly explored or considered in bloodstream infection epidemiology (58). The clear higher BSI rate in men, apart from for *E. coli* infections in those aged 15-40, contrasts with the lack of clear sex effect in many of the resistance trends. The higher BSI levels in women aged 15-40 has been seen previously (59,60) and could reflect the higher urinary tract infection incidence in women (61) which are a common BSI source (62,63).

In addition to the direct implications of our findings on public health policy, understanding of the links between demographics and AMR will be foundational to a deeper understanding of acquisition routes of AMR. In our analysis we explored demographic trends across populations but are limited in our ability to understand the mechanisms behind these trends, where further research is required. One potential avenue for such research is to explore the source of the bacterium causing the bloodstream infection: endogenous following long-term carriage or recent transmission. Age- and sex-related patterns in bloodstream infection source will be influenced by many factors, such as levels of contact with healthcare systems (e.g. hospital stays, previous antibiotic prescriptions (55)), individual behaviour (e.g. causes for hospital admission, rate of contact with other individuals) and inherent biology (e.g. immunosenescence, likelihood of source being a urinary tract or wound infection), as well as varying by bacteria-antibiotic combination. Whilst the contribution of some of these factors have direct links to incidence by age (e.g. immunosenescence contributes to higher sepsis incidence with age (64)), linking these factors to proportion resistance by age is more complicated.

One theory linking the proportion of resistant infections with age is that older individuals are more likely to have weaker immune systems, and therefore are more likely to develop infections due to bacteria they are already colonised with and then enter the healthcare setting, as compared to younger individuals that would be relatively more likely to acquire a resistant bacterium through a transmission event within a healthcare setting. This would have implications for the proportion resistant by demographic characteristics for given bacteria and could be linked to changes in the microbiome with age (65). One of the most surprising species we detected patterns for was *Acinetobacter* species, with strong age-, sex- and subregion-related trends. This could be explained by young men being more likely to attend hospital for trauma compared to women (66), so if for this demographic population the key route to bloodstream infections is from wound infections due to bacteria (such as *Acinetobacter spp*.(67)) acquired in hospital, this could explain the differences we observed in resistant proportion. Differences in incidence and resistance proportion could also be explained by the demographics of those who travel to areas with higher prevalence of *Acinetobacter* species. Key to understanding the influence of the various factors on AMR is detailed knowledge at an individual level, as well as information on community vs hospital acquisition and antibiotic exposure, which we were unable to determine in this study. However, there are indications that for many bacteria, hospital-acquired infections are likely to have higher resistance (24) so changing contact with healthcare would be an important avenue to explore. Exploiting this demographic link could also be used to tackle and determine the key drivers of known subregional variation in resistance prevalence (41). Our work therefore highlights the need for future research on the mechanisms of age- and sex-related AMR trends. In order to achieve this individual-patient level data, linked across primary and secondary care will be essential.

Our research has several limitations. Firstly, we were unable to account for co-morbidities and other syndromes of individuals, which may impact the age groups that are susceptible to different infections. For example, cystic fibrosis patients are known to be particularly susceptible to infections of *P. aeruginosa* (68), whilst also being correlated with the demographics of patients (69). Not including such aspects may particularly bias our work on the future burden of AMR, as the demographics of the syndromes will likely also change over time (69). This also highlights the need to take syndromes into account when prescribing antibiotics, as well as demographic factors, and to record such information alongside AMR data. Our analysis is only of European data, and not split by community or hospital-onset, and as such may not represent universal trends that could vary in other settings, in particular where demographic and healthcare distributions are substantially different.

In addition, the individuals included in this data set may be biased because of variation in whose samples are sent to be tested for resistance. This variation will depend on demographics and can be influenced by the age of the individual, the severity of infection and previous failed antibiotic use, and testing guidelines, among other factors. Understanding the decisions in sampling made by clinicians and other healthcare professionals is a vital area of future study and may account for some of the local level variation we have identified (57). Upstream of the sampling decision, it may also be influenced by healthcare seeking behaviour, for example women are more likely to seek healthcare than men (70), and this also varies by age and potentially by country. Variation in what within hospital settings samples are taken from (ICU, A&E etc.) may also explain some of the national and sub-national variation we observe and which we report, but need further country-specific information to explore, in our sensitivity analysis. By using data from TESSy, which contains only blood and cerebrospinal fluid isolates, likely representing the most serious types of bacterial infection (71) where the vast majority of infections will be hospitalised, these biases should be minimal . However this does not mean they are all sampled, and of those there are many samples will test negatively for infection (72).

In terms of the analysis and modelling in this paper, we chose to limit ourselves to a ‘one-size-fits-all’ approach, applying the same models to each bacteria-antibiotic combination. There is potential for models that are a better fit to the data for specific bacteria-antibiotic combinations, however this approach allowed us to compare model outputs across bacteria-antibiotic combinations, as well as reducing the complexity required. Lastly, we did not attempt to link AMR prevalence with mortality rates. This is because we did not have appropriate information to do this, with age-specific mortality rates and the impact of resistance on infection being hard to estimate, with variations in baselines used (e.g. associated vs attributable (49)). Recent estimates have found that data scarcity makes estimating relative risks of mortality by sub-groups or geographical setting difficult (18).

Future work estimating the burden of AMR and impact of interventions will need to account for these trends by age and sex to accurately capture burden. The complexity in trends in resistance prevalence by age and sex interact with the exponential increase in BSI incidence with age to mean that often, the elderly population, especially men, would still be expected to suffer more infections with resistant bacteria. How this collides with the global shift to older populations (2) and the impact this will have on public health burden as well as AMR spread should be a research priority.

In 2018, the WHO asked “Is the impact of AMR the same for everyone? Do any groups in society face greater or different risks of exposure to AMR or more challenges in accessing, using and benefiting from the information, services and solutions to tackle AMR? If yes, who, why and what can we do about it?” (27). In this paper we go some way to addressing these questions by quantifying how AMR prevalence in bloodstream infections across Europe varies by age and sex, as well as identifying variation at the local level. We show the substantial impacts of including age and sex on AMR, and therefore encourage their inclusion in future data collection and research studies, to improve health outcomes across the spectrum of AMR.

## Data Availability

Patient level data is available upon request from the European Antimicrobial Resistance Surveillance Network (EARS-Net) from the Surveillance System (TESSy) for those who meet the criteria for access to confidential data. https://www.ecdc.europa.eu/en/publications-data/european-surveillance-system-tessy.

## Acknowledgements

We are grateful for all the work done by the staff of the participating clinical microbiology laboratories and of the national healthcare services that provided data to EARS-Net. We also thank Dr Sam Abbot. Dr David Hodgson and Dr Tim Russel for discussing model fitting complexities with us.

The views and opinions of the authors expressed herein do not necessarily state or reflect those of ECDC. The accuracy of the authors’ statistical analysis and the findings they report are not the responsibility of ECDC. ECDC is not responsible for conclusions or opinions drawn from the data provided. ECDC is not responsible for the correctness of the data and for data management, data merging and data collation after provision of the data. ECDC shall not be held liable for improper or incorrect use of the data.

## Appendix S1: How demographic factors matter for antimicrobial resistance – quantification of the patterns and impact of variation in prevalence of resistance by age and sex

### 1. Abbreviations

ACISPP: Acinetobacter spp
ENCFAE: Enterococcus faecalis
ENCFAI: Enterococcus faecium
ESCCOL: Escherichia coli
KLEPNE: Klebsiella pneumoniae
PSEAER: Pseudomonas aeruginosa
STAAUR: Staphylococcus aureus
STRPNE: Streptococcus pneumoniae
#K: number of thousands
AMR: antimicrobial resistance
BSI: bloodstream infection
EARS-NET: European Antimicrobial Resistance Surveillance Network (EARS-Net)
ECDC: European Centre for Disease Prevention and Control
GLASS: Global Antimicrobial Resistance and Use Surveillance System
IQR: interquartile range
MRSA: methicillin resistant S. aureus
WHO: World Health Organisation

### 2. Data cleaning

#### Original data

The original data from the ECDC included 3,549,617 isolates with age, reporting country and different combinations of test results to a variety of antibiotics. This grouped into 510,816 unique combinations of year, age, pathogen, gender, antibiotic tested and country with a total of 9,855,100 antibiotic testing results (either susceptible “0” or resistant “1”) over 46 bacteria-antibiotic groupings. Samples were tested to a different combination of antibiotics: on average 2.7 antibiotic test results were conducted per sample. There were slightly fewer females in the data: 1,445,370 (45%) of isolates were from females.

The proportion of isolates in each age group (0-4, 5-19, 20-64, 65+) and by patient sex are reported in EARS-Net country summaries and on the ECDC Dashboard (e.g. of the 2019 report (1)).

#### Data overview

(1) Laboratory code: There were 1755 unique laboratory codes in the data with a wide range of number of samples per laboratory: an average of 1803 [IQR: 47 - 2202]. No specimens were missing a laboratory code.
(2) Antibiotics: Across the 24 different antibiotic resistance groupings (e.g. some to individual antibiotics, some to families such as macrolides, some to multiple resistance) over the whole dataset, most had more than 50,000 results (susceptible or resistant value), but three were outliers with fewer than 20,000.
(3) Some antibiotic classifications were not to single antibiotics but were summary indicators of “multiple resistances” i.e. in the data a ’1’ indicated multi-resistance when resistance to a set combination of resistances was measured e.g. to macrolides and penicillin. This data was excluded as the aim here was to consider resistance to single antibiotics, not combinations. These ”combined” resistance profiles were generated to look at levels of multi-resistant bacteria in the EARS-Net analysis (e.g. Table 3 in (1)).
(4) Bacteria: Of the 7 bacteria in the dataset, all had >150,000 results apart from *Acinetobacter* species which had ∼50,000. All bacteria were kept in the data.
(5) Countries: Across countries, four countries had approximately a million isolates. There was a substantial decline across the other countries. 5 countries had fewer than 12,000 isolates. We did not exclude any country from the analysis.
(6) Years: Across the 19 years (2002–2020), the earlier time points had fewer data (∼65K results) compared to more than ∼750K from 2016. This was distributed differently across different pathogens and so we did not explicitly exclude certain years. Instead, we focused our analysis on the final five years prior to the COVID-19 pandemic i.e. 2015-2019. Subsetting the data to this time period only removed the country Lithuania which had no linked gender data for this time period.

#### Missing data distributions

To determine if the missingness in the data was completely at random, we investigated the missingness by key variables (age, sex and susceptibility). We considered the pattern overall as we’d expect different denominator patterns for each bacteria-antibiotic, but also explored the proportion of missingness for each bacterial species and for the case study of *S. aureus*.

Over the whole dataset, there were a total of 1,902,271 individual records over the 2015-2019 time period, of which 6.0% were missing age only, 1.5% were missing sex only and 2.2% were missing both age and sex. To check for biases in missingness we compared the distribution of ages with those missing sex and those not missing sex (Figure 1A). We also compared the distribution of sex with those missing and not missing age (Figure 1B). In both cases differences were minimal. As a case study, we ran the same analysis on just the *S. aureus* patients (Figure 2). To further confirm the minimality of differences a logistic regression was performed on those with missing gender information compared to those without missing information on the *S. aureus* subset, however these models did not reach convergence. This was also the case when looking at missingness in age. Therefore, as we could not investigate this further and the crude comparison showed minimal differences by missingness, imputation of missing data was not performed.

**Figure 1:**
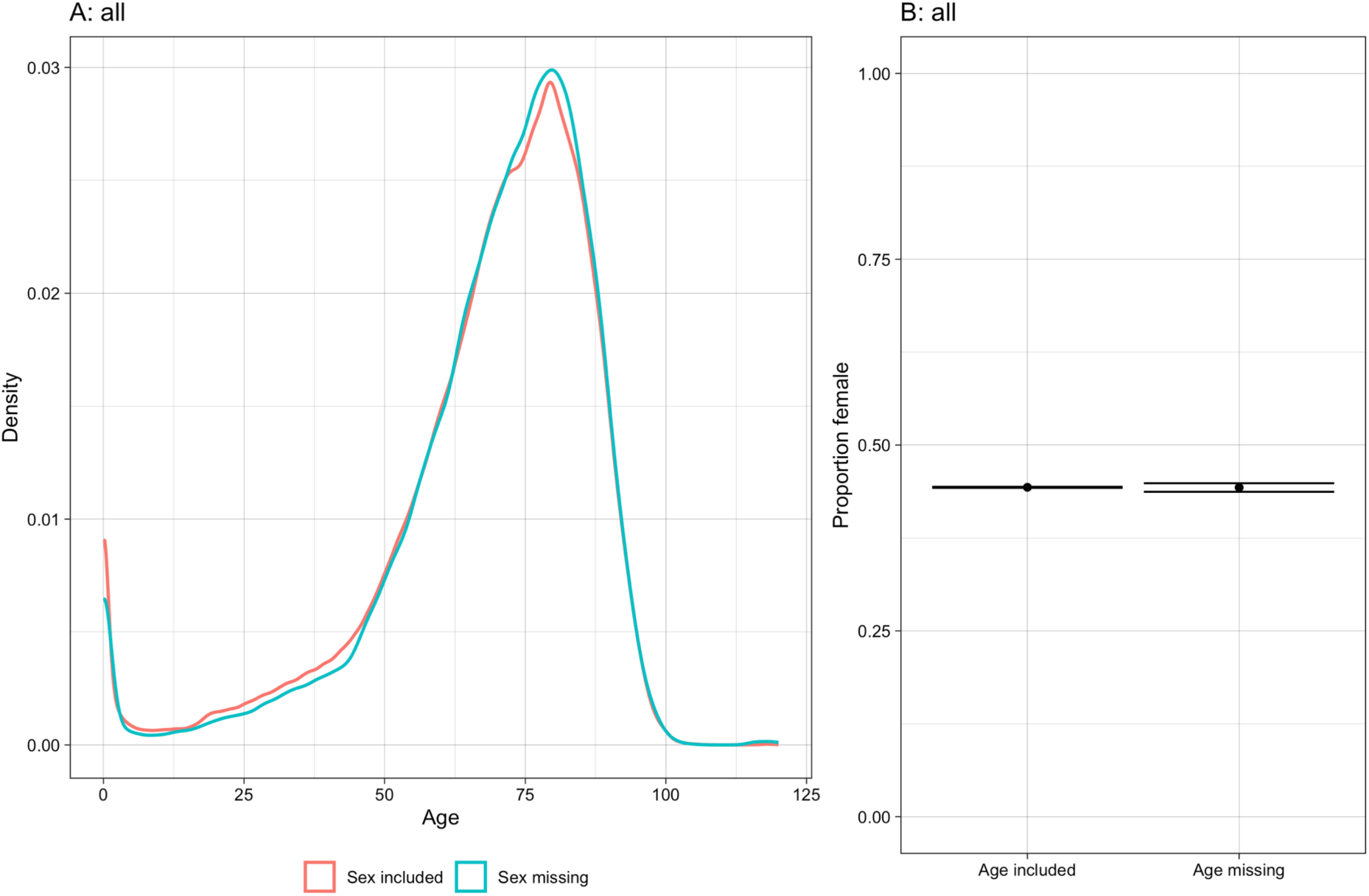
A) Density of individuals across age groups, for those individuals including and missing sex. B) Proportion and 95% CI of female indivduals, subsetted for those including and missing age.

**Figure 2:**
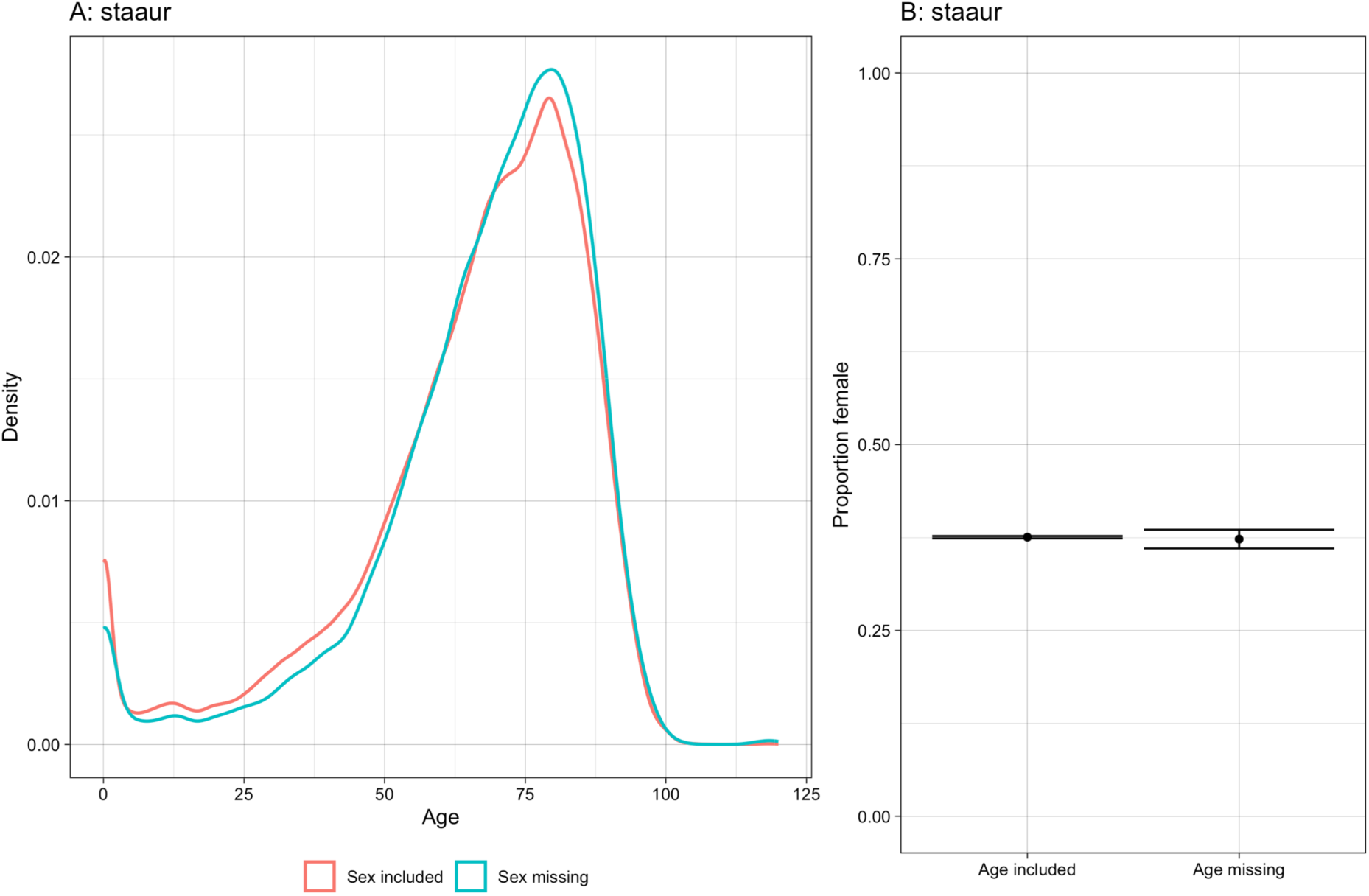
A) Density of individuals across age groups infected with S.aureus, for those individuals including and missing sex. B) Proportion and 95% CI of female indivduals infected with S.aureus, subsetted for those including and missing age.

Not all samples were tested for each resistance type, and hence were coded as NA values in the data. The proportion of samples that were not tested for susceptibility ranged between 1.4% and 51.8%, depending on the bacteria-antibiotic combination (Table 1).

**Table 1:**
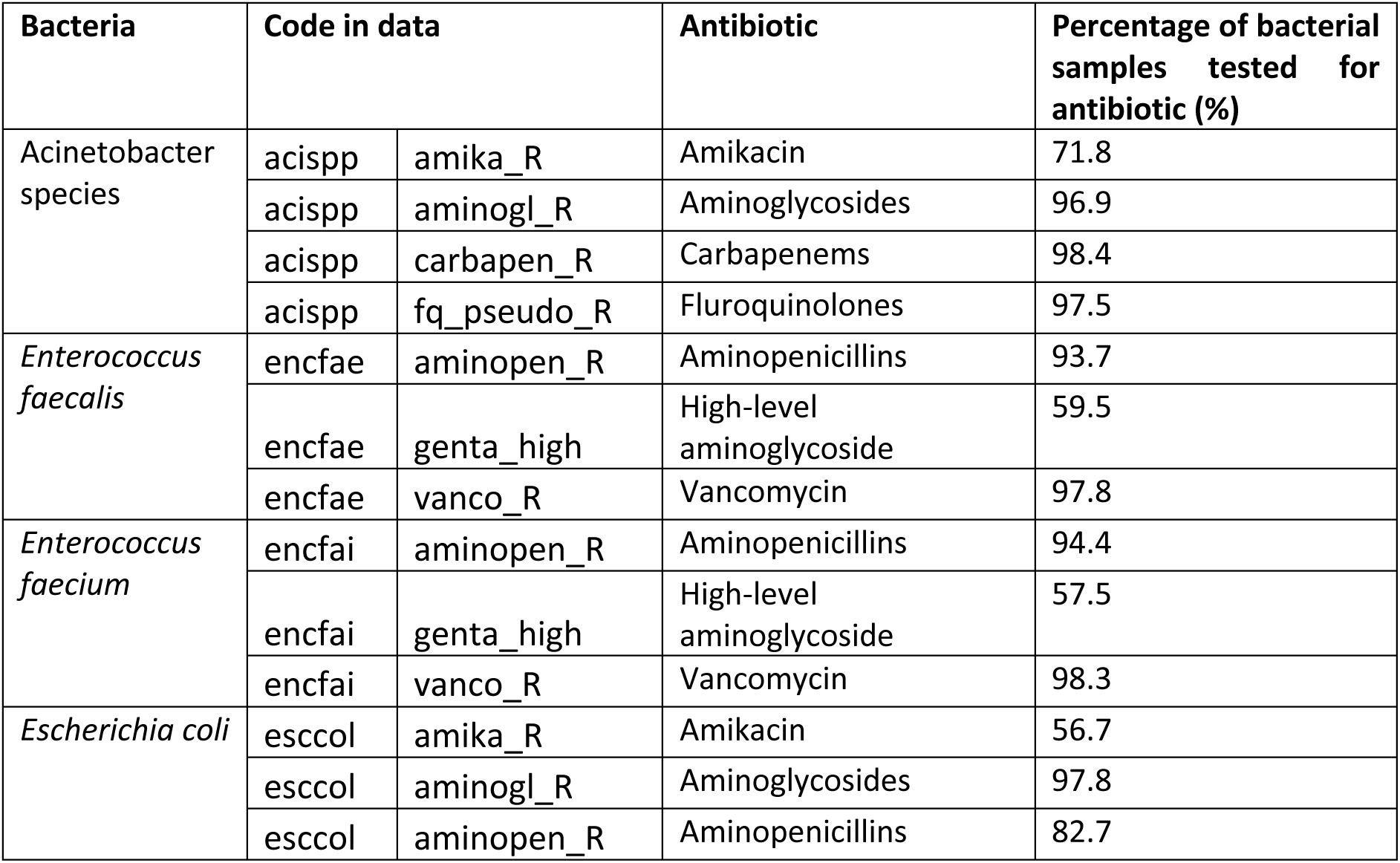
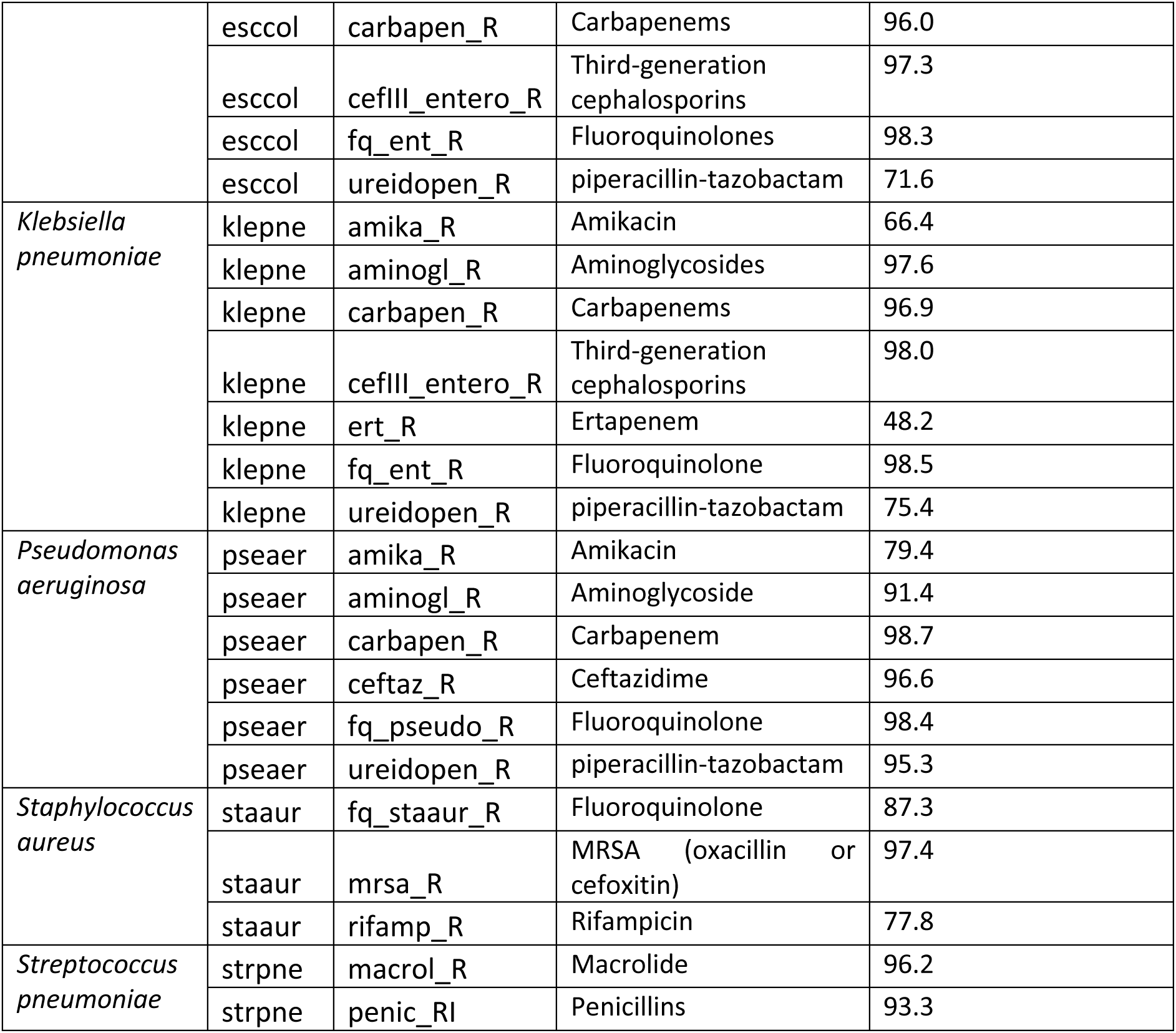
Percentage of bacterial samples tested for resistance by bacteria and antibiotic.

We analysed the missingness of susceptibility test data for our case study MRSA, by age and sex. The distribution of ages between those missing and not missing susceptibility information followed the same trend and there were no differences in susceptibility missingness by sex (Figure 3).

**Figure 3:**
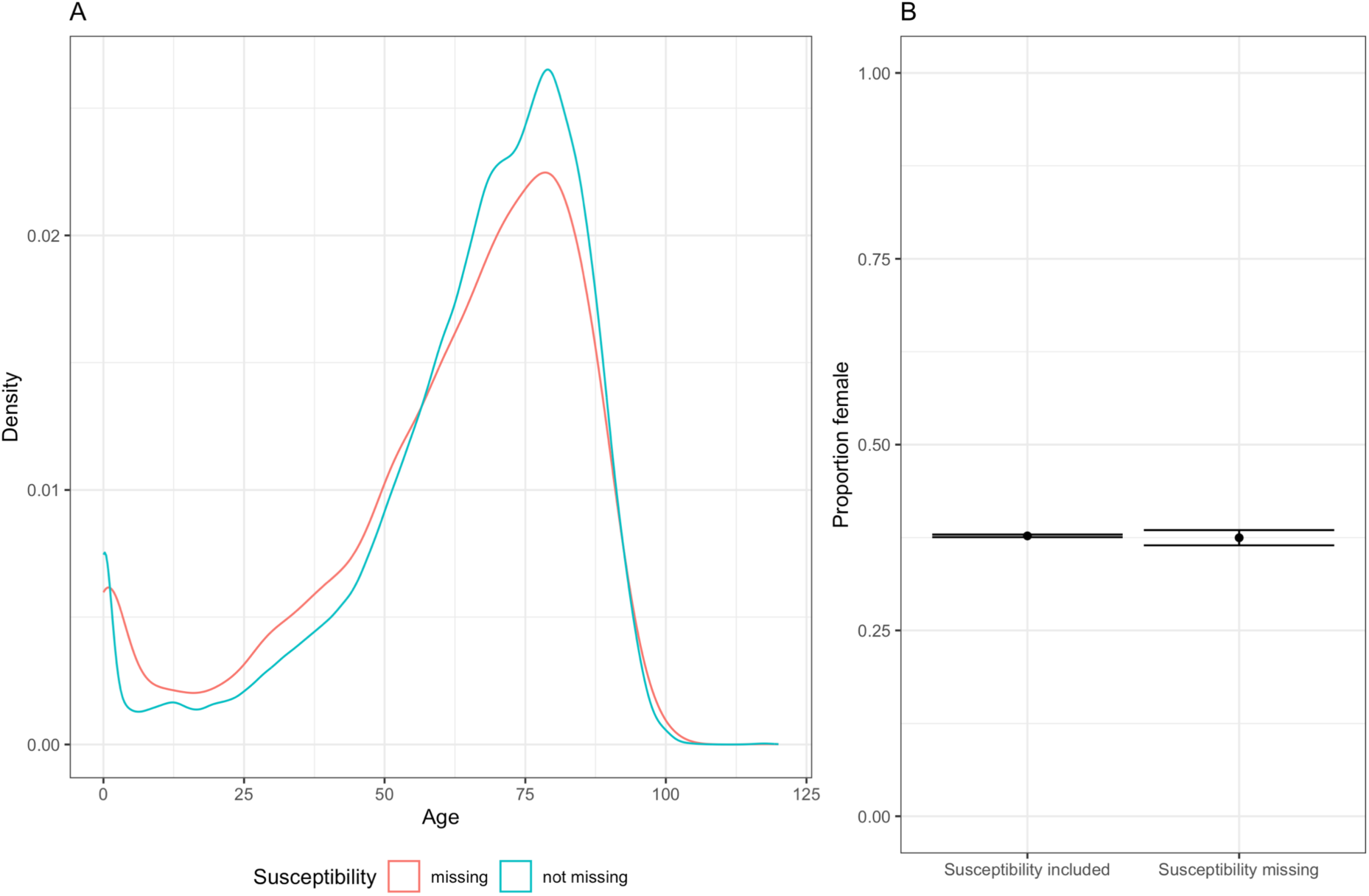
A) Density of individuals across age groups infected with S.aureus, for those individuals including and missing susceptibility tests. B) Proportion and 95% CI of female indivduals infected with S.aureus, subsetted for those including and missing susceptbility tests.

#### Final cleaned dataset

The final dataset filtered for data with (a) age and sex values, (b) for the time period 2015-2019, with (c) multi-resistance class groupings removed and (d) only those aged 1 or older, consisted of data on 17 bug-drug combinations across 8 bacteria (Table 2) and 29 countries (Table 3). Our main analysis therefore used a dataset consisting of a total of 3,881,140 measures of susceptibility consisting of 40% of the original dataset. The average proportion resistant was 30% (21% standard deviation) (across single year age and bug-drug combinations).

**Table 2:**
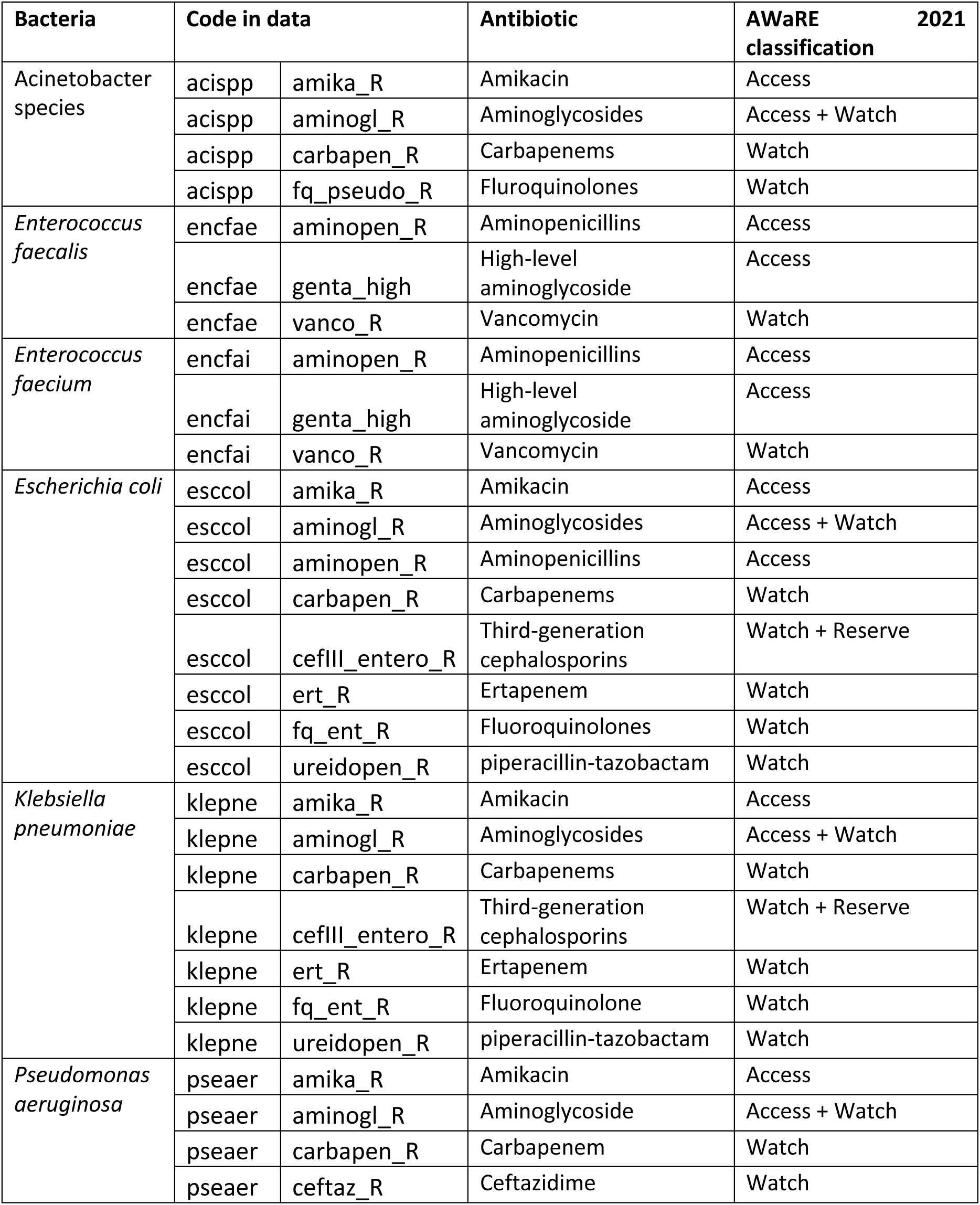
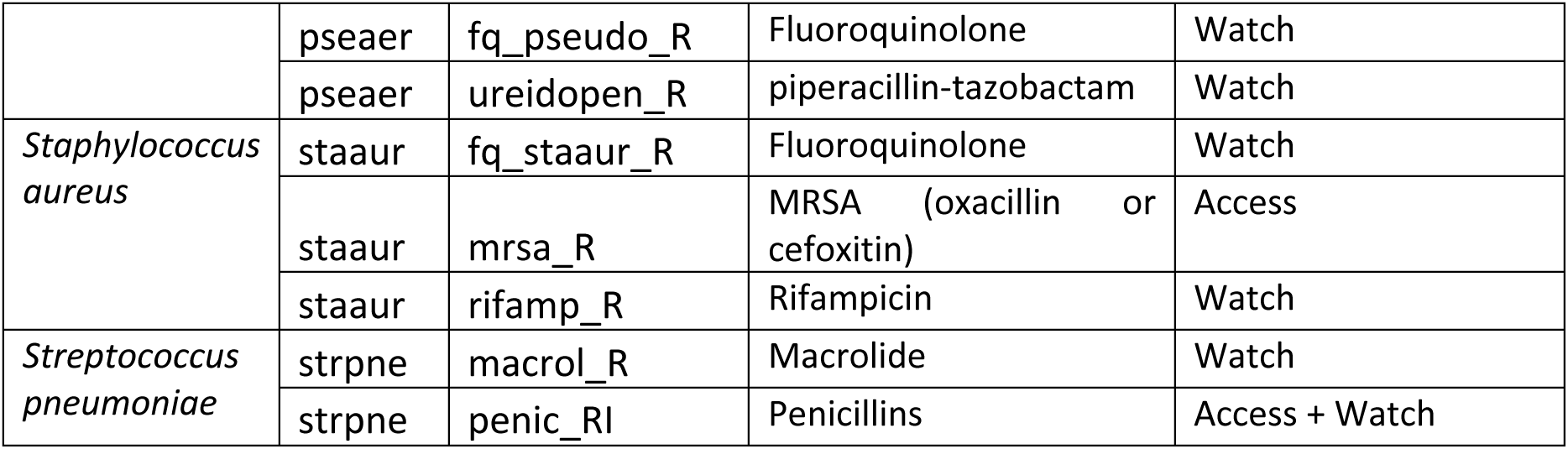
AWaRE classification of bacteria antibiotics in data set.

| Bacteria | Code in data |  | Antibiotic | AWaRE 2021 classification |
| --- | --- | --- | --- | --- |
| Acinetobacter species | acispp | amika_R | Amikacin | Access |
|  | acispp | aminogl_R | Aminoglycosides | Access + Watch |
|  | acispp | carbapen_R | Carbapenems | Watch |
|  | acispp | fq_pseudo_R | Fluroquinolones | Watch |
| *Enterococcus faecalis* | encfae | aminopen_R | Aminopenicillins | Access |
|  | encfae | genta_high | High-level aminoglycoside | Access |
|  | encfae | vanco_R | Vancomycin | Watch |
| *Enterococcus faecium* | encfai | aminopen_R | Aminopenicillins | Access |
|  | encfai | genta_high | High-level aminoglycoside | Access |
|  | encfai | vanco_R | Vancomycin | Watch |
| *Escherichia coli* | esccol | amika_R | Amikacin | Access |
|  | esccol | aminogl_R | Aminoglycosides | Access + Watch |
|  | esccol | aminopen_R | Aminopenicillins | Access |
|  | esccol | carbapen_R | Carbapenems | Watch |
|  | esccol | cefIII_entero_R | Third-generation cephalosporins | Watch + Reserve |
|  | esccol | ert_R | Ertapenem | Watch |
|  | esccol | fq_ent_R | Fluroquinolones | Watch |
|  | esccol | ureidopen_R | piperacillin-tazobactam | Watch |
| *Klebsiella pneumoniae* | klepne | amika_R | Amikacin | Access |
|  | klepne | aminogl_R | Aminoglycosides | Access + Watch |
|  | klepne | carbapen_R | Carbapenems | Watch |
|  | klepne | cefIII_entero_R | Third-generation cephalosporins | Watch + Reserve |
|  | klepne | ert_R | Ertapenem | Watch |
|  | klepne | fq_ent_R | Fluroquinolone | Watch |
|  | klepne | ureidopen_R | piperacillin-tazobactam | Watch |
| *Pseudomonas aeruginosa* | pseaer | amika_R | Amikacin | Access |
|  | pseaer | aminogl_R | Aminoglycoside | Access + Watch |
|  | pseaer | carbapen_R | Carbapenem | Watch |
|  | pseaer | ceftaz_R | Ceftazidime | Watch |
|  | pseaer | fq_pseudo_R | Fluoroquinolone | Watch |
|  | pseaer | ureidopen_R | piperacillin-tazobactam | Watch |
| *Staphylococcus aureus* | staur | fq_staur_R | Fluoroquinolone | Watch |
|  | staur | mrsa_R | MRSA (oxacillin or cefoxitin) | Access |
|  | staur | rifamp_R | Rifampicin | Watch |
| *Streptococcus pneumoniae* | strpne | macrol_R | Macrolide | Watch |
|  | strpne | penic_RI | Penicillins | Access + Watch |

**Table 3:**
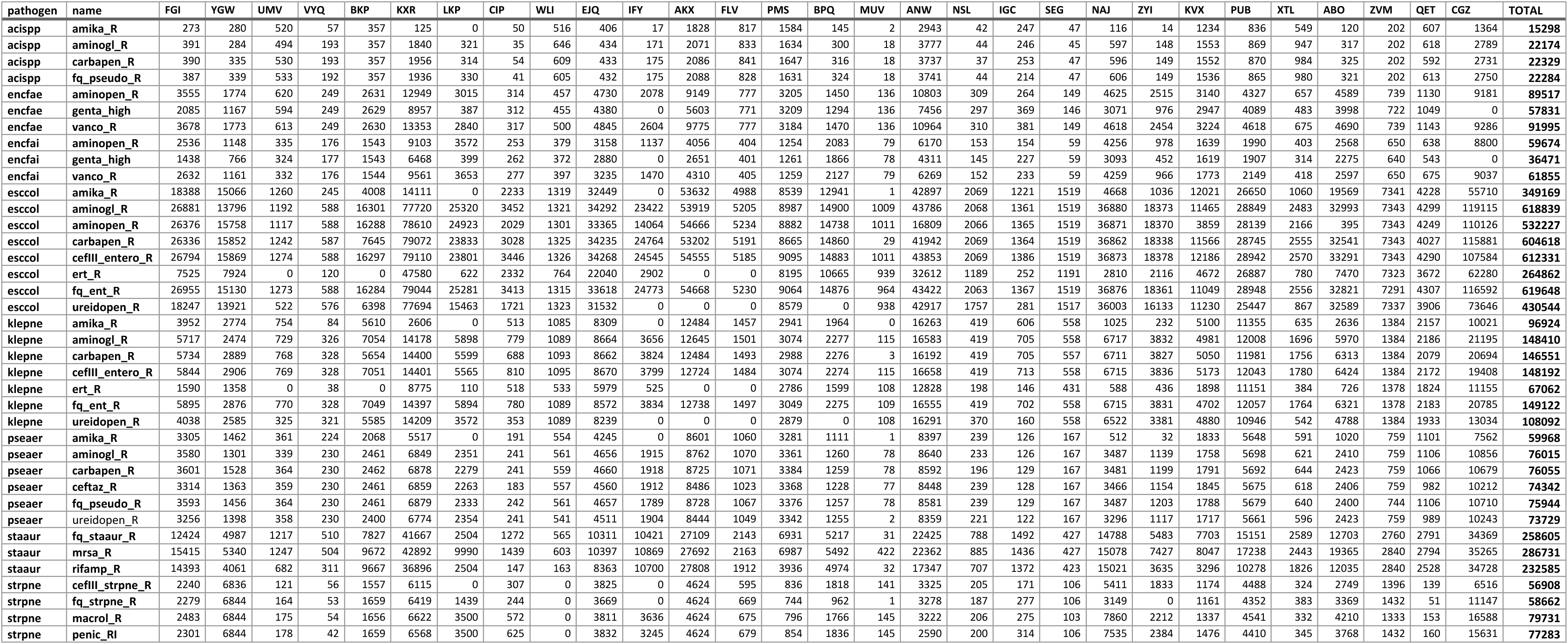
Number of susceptibility samples by bacteria-antibiotic (rows) and country (column) in cleaned dataset. Country labels are random anonymised three letter code used for this analysis only.

### 3. Incidence calculations

The calculation of incidence used all data that had (a) age and (b) sex (male / female) recorded. Excluding those isolates without this information removed 4.8% or 7.1% of isolates respectively. Due to the overlap in missing data, the final data set included 91% of the original isolates (a total of 3,231,153 isolates).

The raw data can be used to calculate the number of *isolates* taken from patients of each age which can be standardised by the population in each age group. However, the network of contributing laboratories is not a population-based surveillance network – the number of isolates does not always reflect the total number of infections in a country. Therefore, in order to estimate the incidence of bloodstream infections by age we need to inflate the reported numbers by some indication of what proportion of the infections are captured i.e. the coverage.

#### Coverage complexity

The hurdles to understanding coverage are two-fold. Firstly, not all microbiology laboratories responsible for processing isolates are linked to the EARSS/EARS-Net network. In the ECDC AMR report of 2017 (2) (and previous years), the number of laboratories reporting at least one isolate to the EARSS/EARS-Net surveillance dataset is shown for the 2000-2016 period. The number of laboratories reporting is stable or increasing over this period, showing how coverage has improved.

However, the size and number of laboratories varies by country and hence it is hard to get an estimate for coverage or representativeness from this. Moreover, the overlapping hospital population catchment areas and movement of patients for care seeking means that the exact proportion of the population included in the data is highly difficult to assess. It requires local knowledge and will likely always be an approximation.

Hence, ECDC AMR reports since 2018 have asked countries to report an “Estimated population country coverage”. This is a self-assessed national coverage and sample representativeness value as estimated by the National Focal Points for AMR and/or Operational Contact Points for AMR (Table 2.1 in the 2018 report, Table 1 in the 2019 report, Table 2 in the 2020 report (3)). This is reported across all bacteria in the EARS-Net reports and shows a large variation between countries and some variation in time (Figure 4).

**Figure 4:**
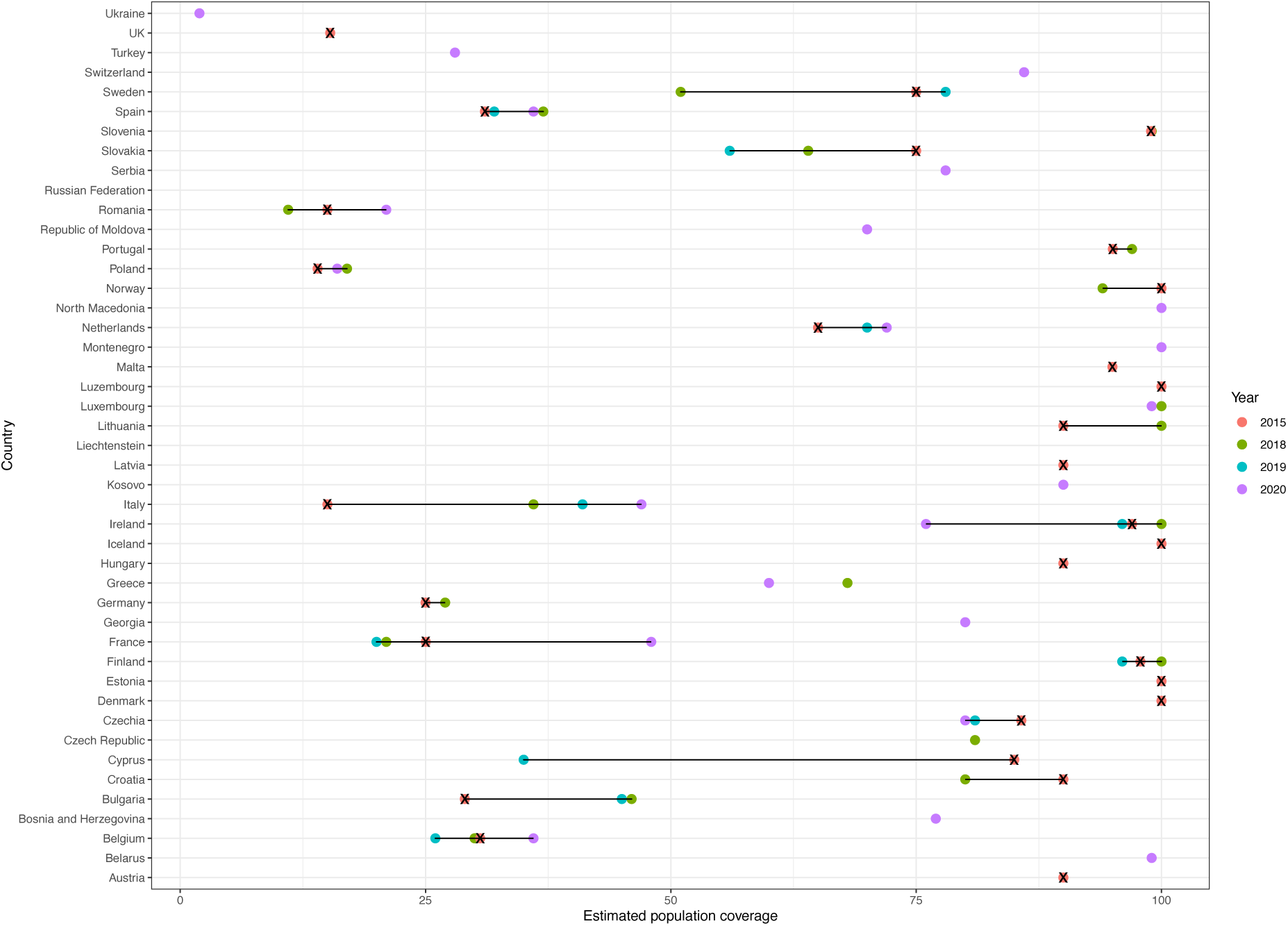
Combined estimated population coverage of the isolates reported in EARS-Net from the annual antimicrobial resistance surveillance reports (3) and estimates reported to Cassini et al. (shown with crosses).

It is likely that there is further variation in coverage or representativeness when comparing between bacteria. This is shown in data collected by Cassini and colleagues for their estimates of deaths and disability-adjusted life-years caused by antibiotic resistance bacteria in 2015 (4). Designated contact points in each Member State were given the possibility of providing an estimated “percentages of population covered” to give national population coverage (Table 3 in the Supplementary material of (4)). This showed that despite wide variation between countries (Figure 5), for 70% (21/30) of countries that reported values, the estimated coverage was the same across bacteria in 2015. Two countries had more than 50% variation between coverage –high coverage of *S. pneumoniae* (67%/87%) being an outlier against 18% coverage of other bacteria or 24% for others apart from *Acinobacter spp.* (8%) for France and Belgium respectively.

**Figure 5:**
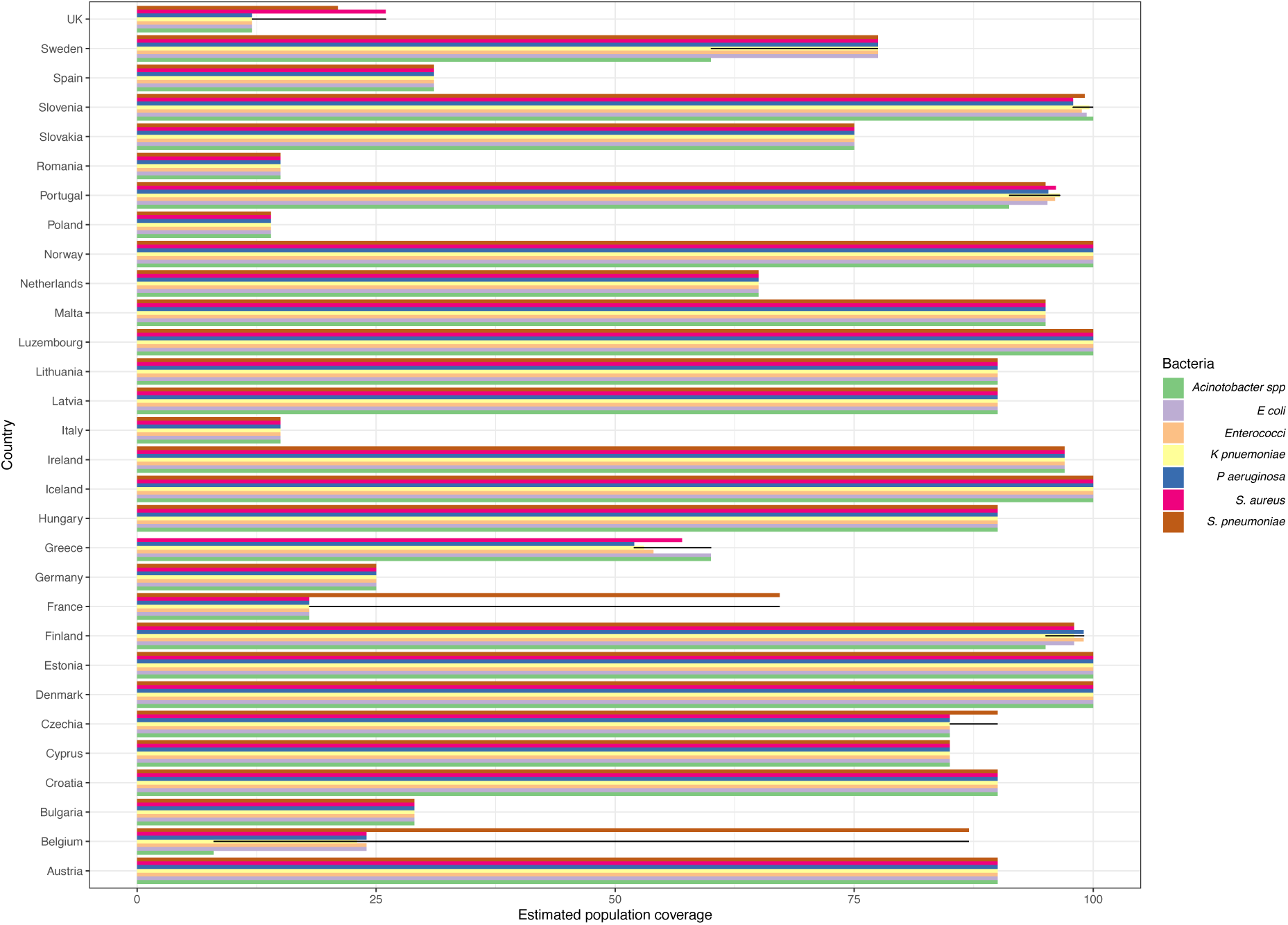
Estimated population coverage of EARS-Net from country experts as reported to the Cassini study (4).

The second hurdle is the variation in local sampling procedures – not just what patients might have their isolates sent to the laboratory but how many patients with an infection will even be sampled and where within the hospital and when. EARS-Net has attempted to address this by considering the blood culture rate (blood culture sets / 1,000 patient-days). This is complicated further by the varying country definitions of a blood culture set as well as a “patient day”. Thus, it is hard to use this indicator – in the ECDC reports it is seen as another factor to bear in mind when interpreting resistance trends and Cassini et al do not use it, after consideration, in their methods.

The added complexity of any variation in sampling by age or patient sex has no information to support any assumptions.

Only the values for Italy (Figure 4) gave a significant trend under a linear model testing for increasing coverage over time, and so it was decided to

(1) use the country level values to inflate number of isolates where they exist for estimates of incidence
(2) to use the last value available for a country as the value to inflate the data for total number of isolates in projections. Both the number of isolates that are likely to be reported to EARS-Net and the inflated totals per country are reported.

Our sensitivity analysis explored using the minimum coverage value instead of the last.

Cerebrospinal fluid isolates represent a small minority of the EARS-NET data (< 4% of all isolates, only rising to 6% for *S. pneumoniae*) and are likely to also have a BSI, so, following Cassini et al, we include them in our estimates of BSI.

### 4. Population trends – 2000 to 2021

The number of individuals in each age and sex group for the period of 2000-2021 was needed for the calculation of age-standardised incidence rates. Population sizes for 5yr age groupings were available from the World Bank DataBank (5) up to the age of 80 and then all adults were pooled. These are World Bank staff estimates using the World Bank’s total population and age/sex distributions of the United Nations Population Division’s World Population Prospects: 2019 Revision (6). The trends vary substantially by country (Figure 6) with an increase in the proportion in older ages present in most countries (Figure 7).

**Figure 6:**
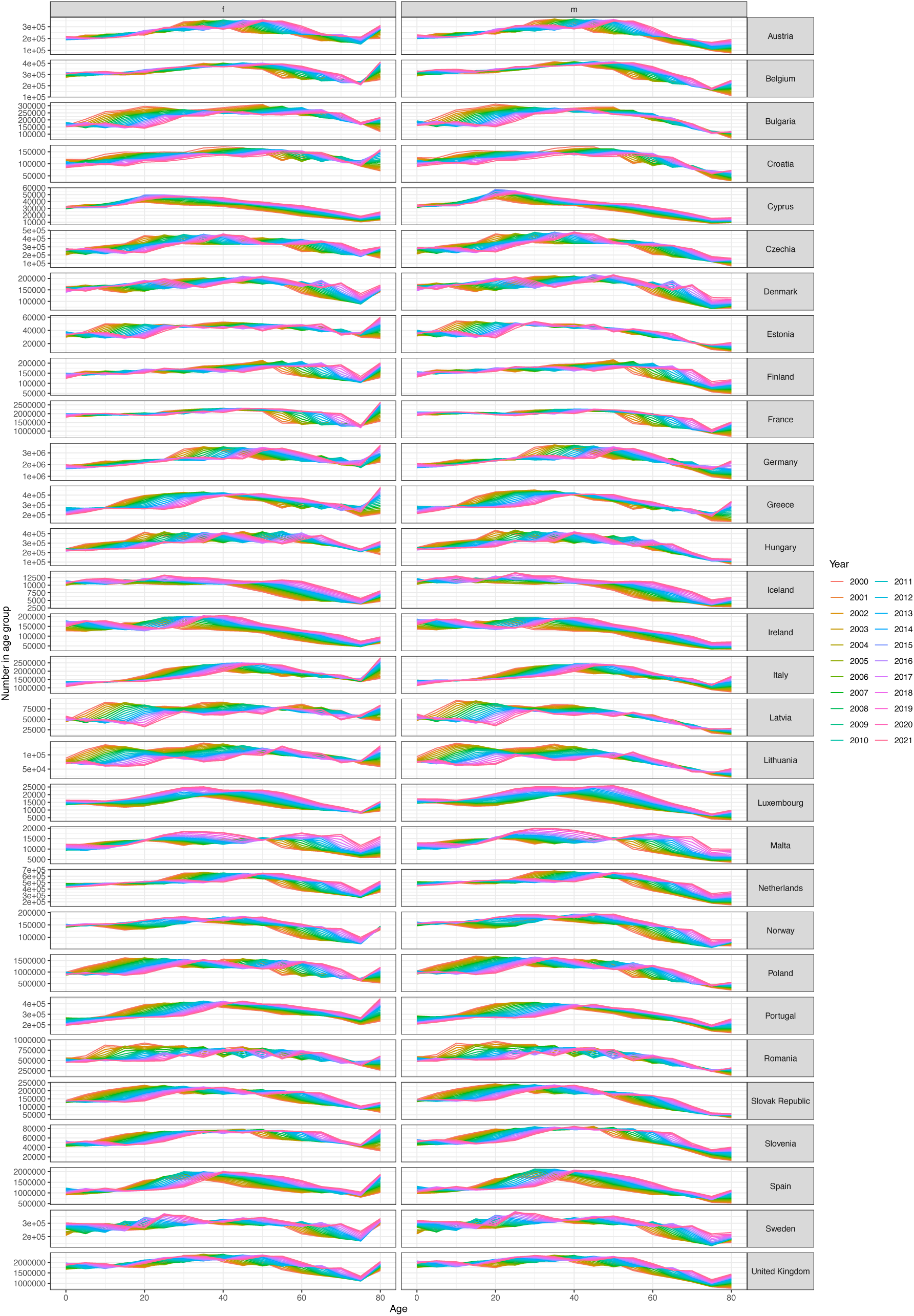
Population sizes by age group (5yr age bands represented by lowest age, x axis, e.g. plotted at age 0 for age band 0-5yo), sex (columns, f = female, m = male) and country (rows) for each year (colour) from the World Bank.

**Figure 7:**
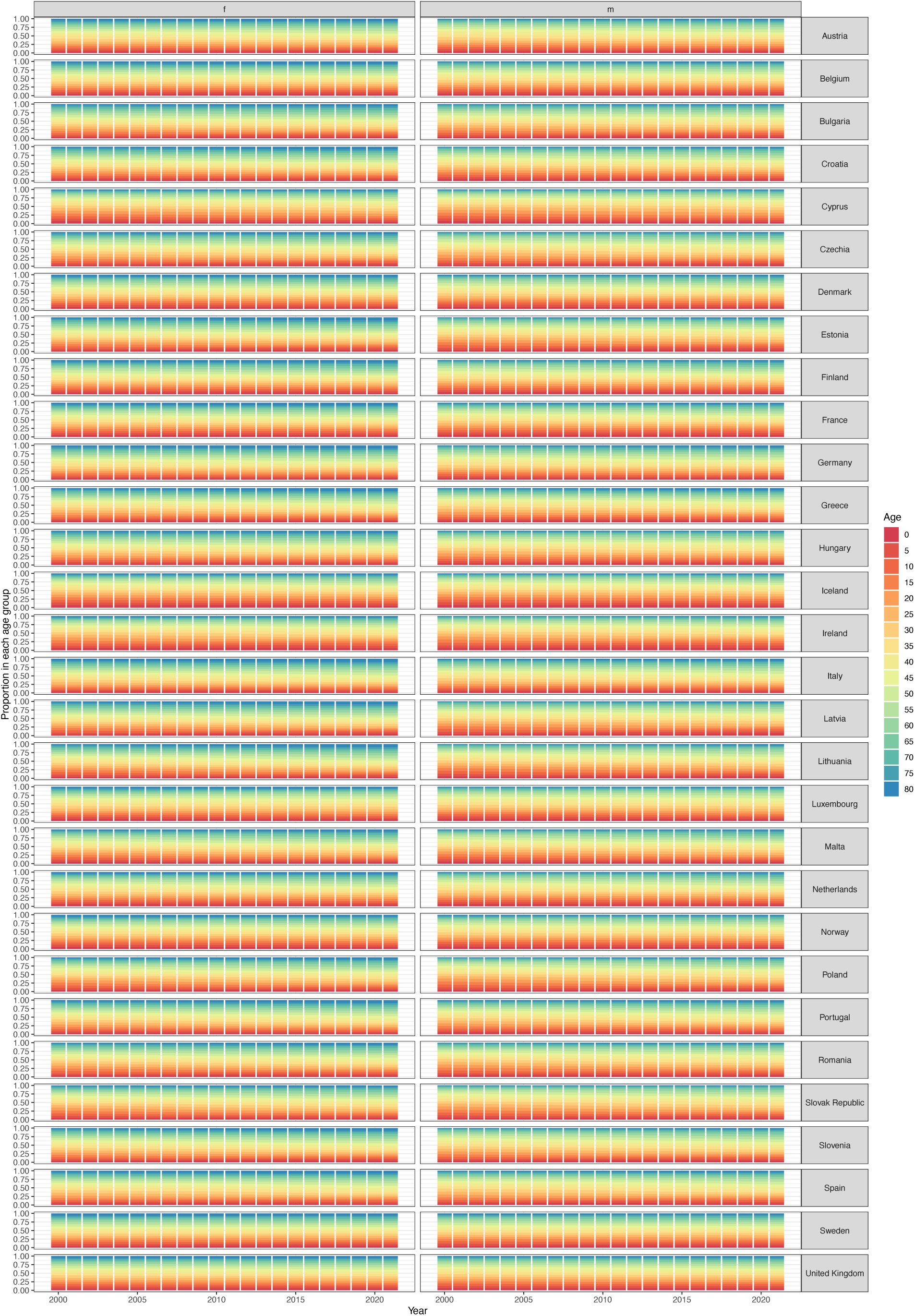
Proportion of population (y axis) in each age band (colour) over time (x axis) for each country (row) and each sex (column) from the World Bank.

### 5. Model fitting

#### Variables included

To determine whether country and laboratory level variation should be included in the model, we ran a variance-components model (using the R *glmer* functions) for MRSA, using this as a case study for the other bacteria-antibiotic combinations. Results showed that 33% of susceptibility variation lies between countries, 38% of susceptibility variations lies between laboratory ID and country combined. Figure 8 shows the variation across different laboratories and countries.

**Figure 8:**
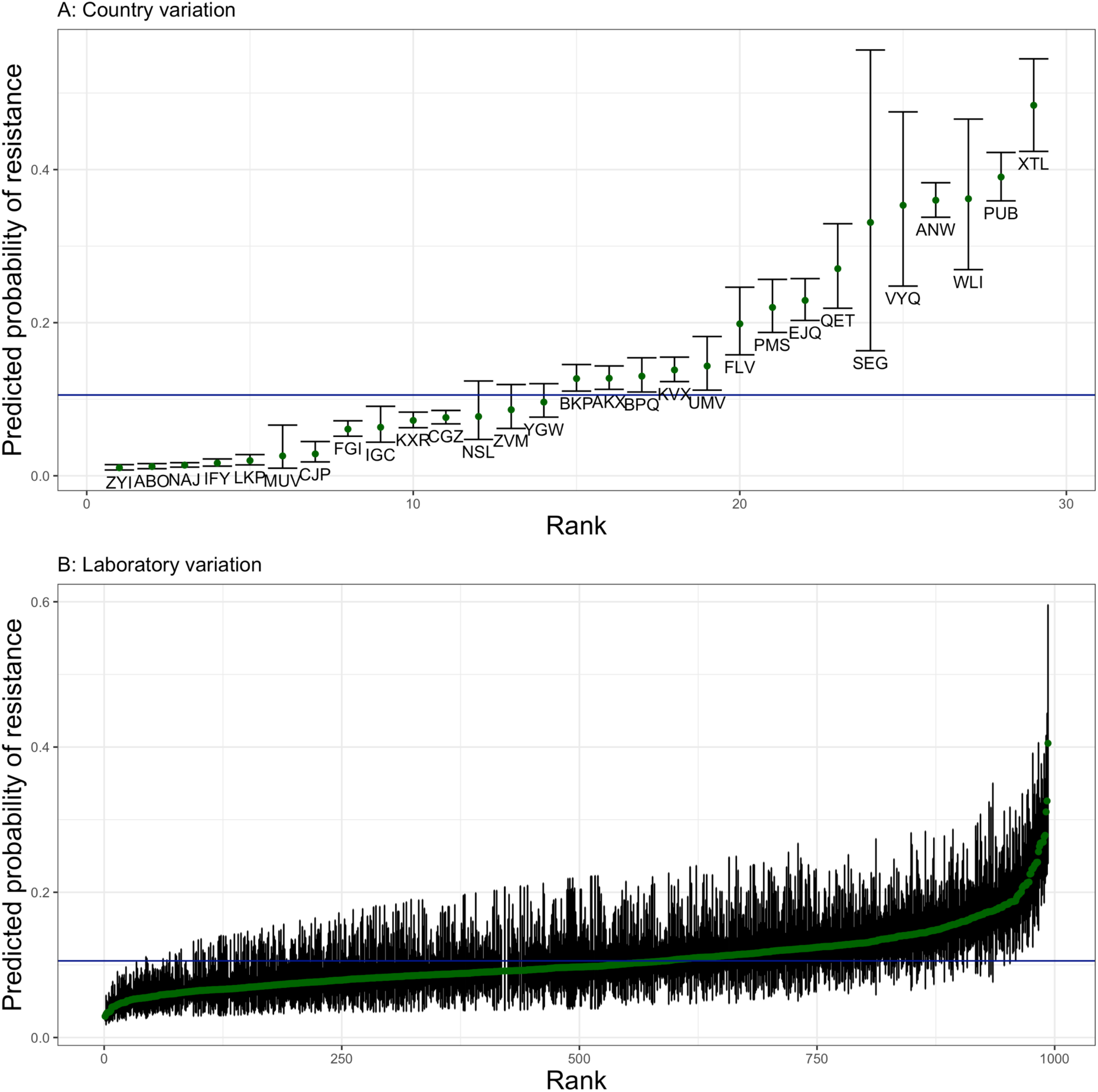
Predicted probability of resistance across (A) hospitals within a country and (B) laboratory ids over all countries.

#### Variable transformations

To achieve efficient model convergence, we transformed the input variables in our model to be on a similar scale. The transformations were as follows:

Model age <-Age / 100

Model age^2^ <-Model age * Model age

Years were converted onto a scale of 0-1 corresponding to the years 2015-2019.

#### Fitting algorithm

We used the brms package in R to run our models using stan software (7). The algorithm used was no U-turn sampling (NUTS) which we ran for 3000 iterations for each model. We deemed models to not have converged if more than 1 divergent transition occurred. Bacteria-antibiotic combinations that did not converge were: *Escherichia coli* – Ertapenem*, Enterococcus faecalis* – aminopenicillins and *Enterococcus faecalis* – vancomycin. We therefore excluded these combinations from our analysis, as they did not converge and they were not considered key bacteria-antibiotic combinations: they were not included in recent ECDC analysis reports (e.g. (8).

## Appendix S2: How demographic factors matter for antimicrobial resistance – quantification of the patterns and impact of variation in prevalence of resistance by age and sex

### 1. Additional results: subregion analysis

We observed substantial variation by subregion, matching previously reported large variation in resistance prevalence (with a north-to-south and west-to-east gradient of resistance) (1)) (Figure 1). Southern and Eastern generally had higher resistance prevalence with substantial age / gender differences in south + east especially for *Acinetobacter spp.* and MRSA patterns driven by south/east Europe, whilst for others the differences are less important e.g. for aminopenicillin resistance in *E. coli* (Figure 2). The patterns are still more similar within a bug-drug than across subregions: i.e. the same across subregions and then different between bug-drugs.

**Figure 1:**
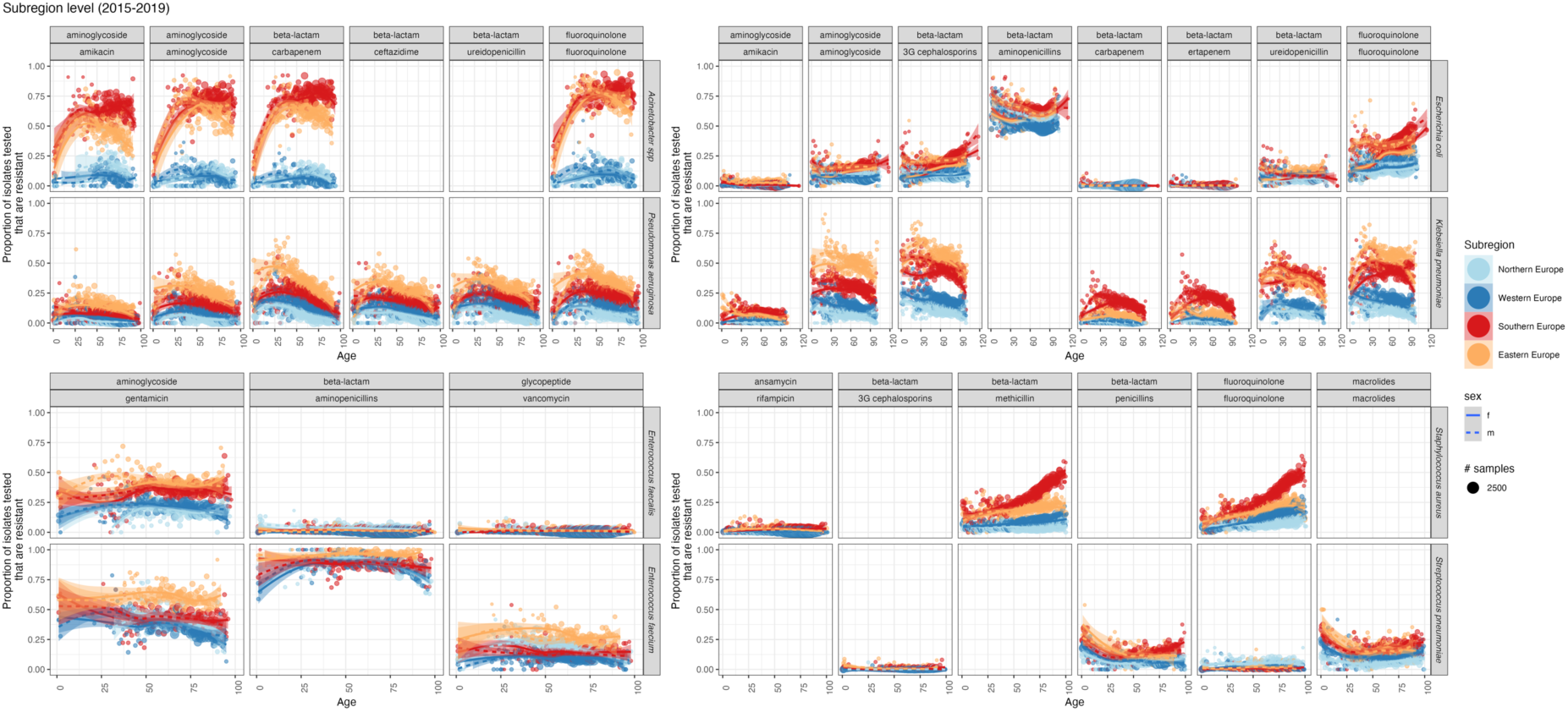
Trends in resistance prevalence vary by antibiotic, bacteria and demographic factors across the subregions of Europe. The proportion of isolates tested (y axis) that are resistant to each antibiotic (facet) within drug families (columns and colour) for each bacteria (row) is shown for all European data over 2015-2019 by age (x axis). Data is shown as points with number of samples indicated by size of point. Shaded areas are 95% confidence intervals around the LOESS fit line by gender (linetype and shade).

**Figure 2:**
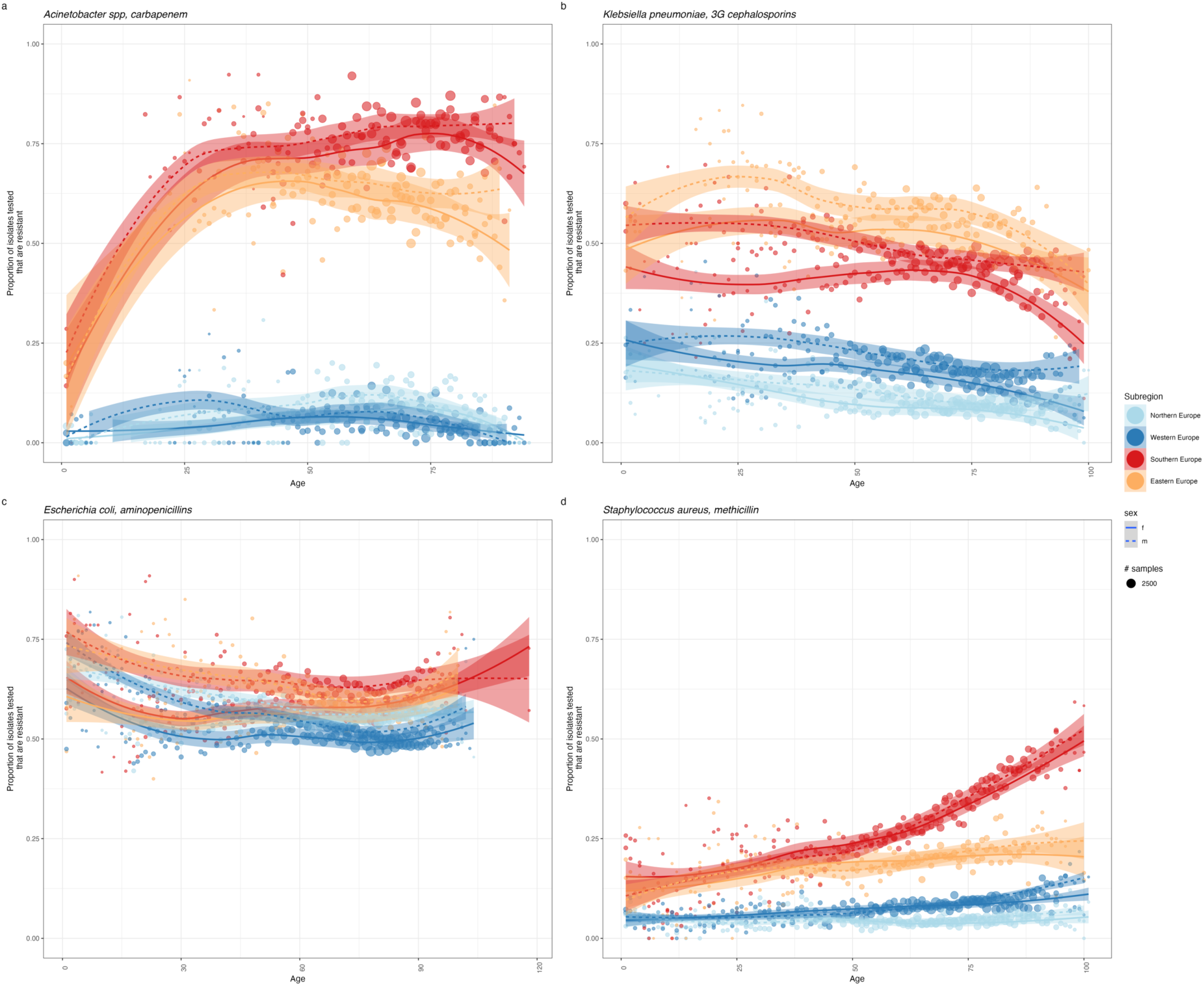
Zoomed in examples of how trends in resistance prevalence vary by antibiotic, bacteria and demographic factors across the subregions of Europe. The proportion of isolates tested (y axis) that are resistant to each antibiotic (facet) within drug families (columns and colour) for each bacteria (row) is shown for all European data over 2015-2019 by age (x axis). Data is shown as points with number of samples indicated by size of point. Shaded areas are 95% confidence intervals around the LOESS fit line by gender (linetype and shade).

### 2. Additional results: Incidence

The incidence of infection was calculated for all 29 countries in both datasets (population size and infection incidence). For this work we only included countries if they had reported data on the estimated coverage of these laboratories for any of the years this data was available: 2015, 2018-2020.

#### Country level data and analysis

These 28 countries were: "Austria", "Belgium", "Bulgaria", "Croatia", "Cyprus", "Czechia", "Denmark", "Estonia", "Finland", "France", "Germany", "Greece", "Hungary", "Iceland", "Ireland", "Italy", "Latvia", "Luxembourg", "Malta", "Netherlands", "Norway", "Poland", "Portugal", "Romania", "Slovak Republic", "Slovenia", "Spain", "Sweden" and “United Kingdom”. Not all countries were included in each data point. There was substantial variation between countries.

What is similar between countries is that there appears to be a trend to increasing incidence rates over time for those with data from 2015-2020. This reflects trends seen in earlier reports for *S. aureus* across Europe from the same data for 2005-2018 (2) and earlier trends for all BSI across Europe for 2002-2008 (3). In individual countries increases have been seen in BSI e.g. due to *E. faecalis* in Switzerland 2013-2018 (4,5), in England and Wales 1990-1998 (6) or England 2004-2008 (7). We report this trend here but do not use it in our analyses.

**Figure 3:**
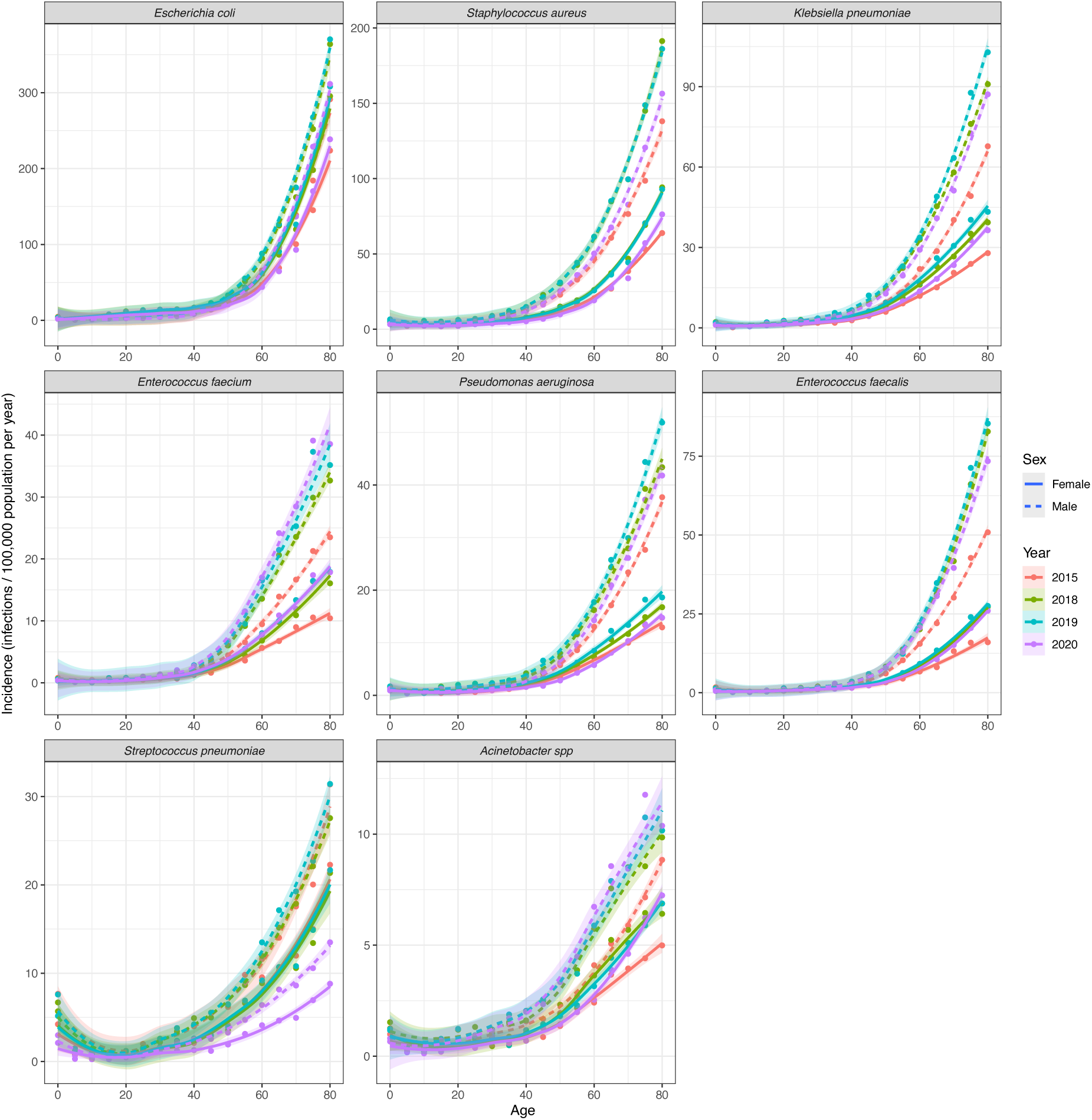
Incidence of bloodstream infections per 100,000 population per year across European countries for 8 bacterial pathogens (panel) for multiple years (colour) and split by sex (solid line = female, dashed line = male). Shaded areas are 95% confidence intervals using a LOESS fit.

### 3. Additional results: total number of resistant infections by age

The combination of exponential increase in infection by age with the proportion resistant, leads to an exponential increase in the number of infections with resistant bacteria by age (Figure 4).

**Figure 4:**
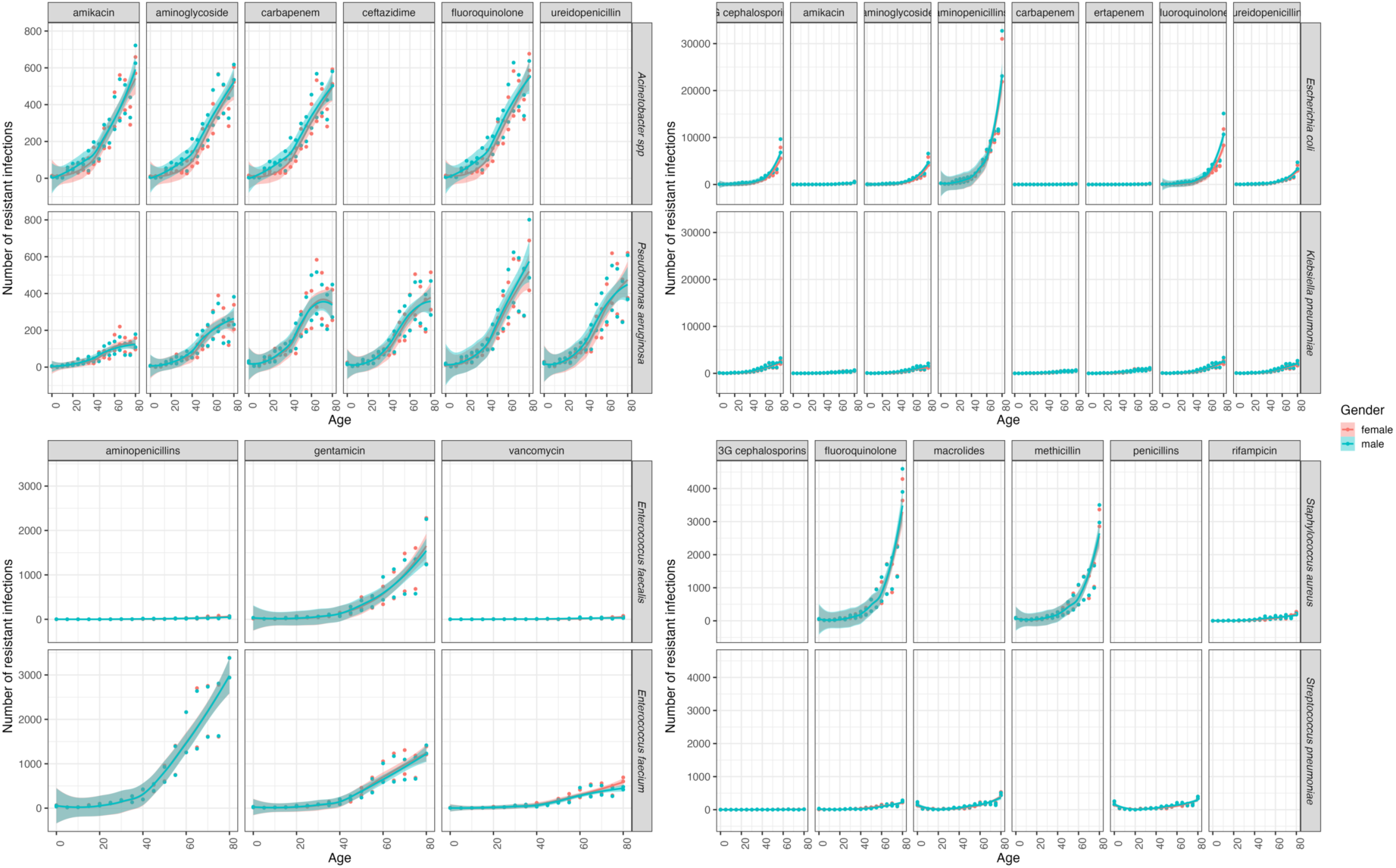
Total number of resistant infections by age in 2019 across Europe

### 4. Additional results: Posterior estimates from model fit

Table 1 shows all the fixed effect parameter values and their 95% posterior estimates.

**Table 1:**
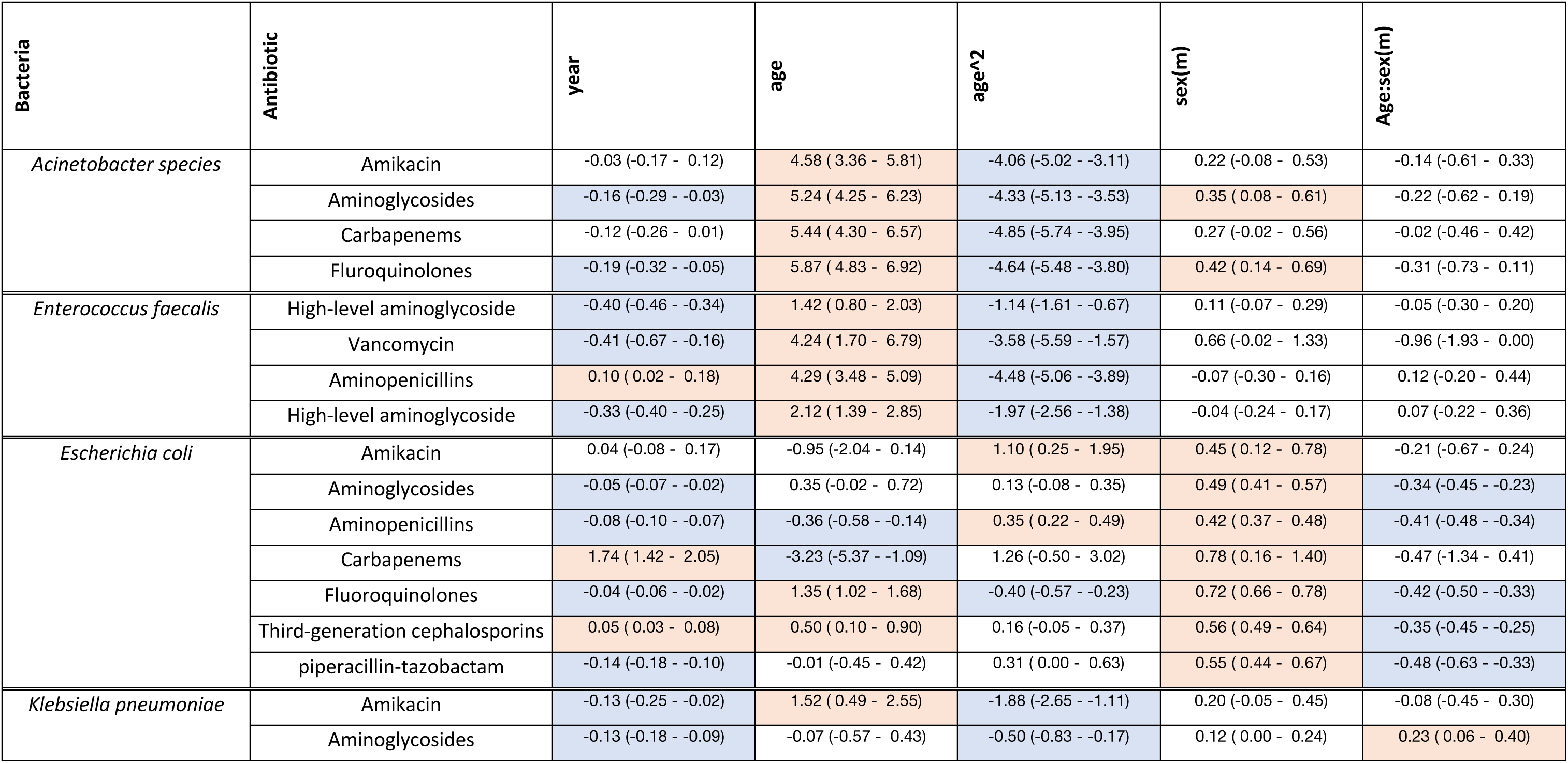
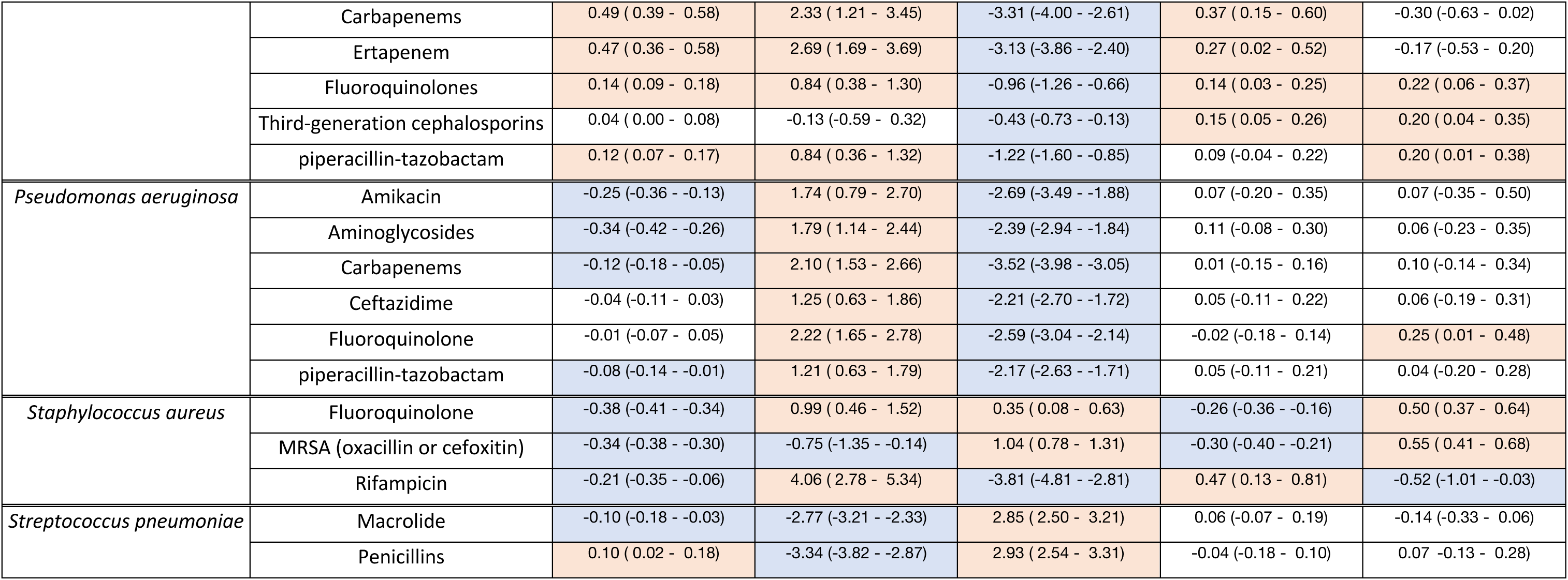
Posterior estimates for fixed effect parameter values. Blue indicates the 95% CI does not cross the null line. Shading matches Table 1 of main paper: orange shading indicates a positive significant effect and blue indicates a significant negative effect.

### 5. Additional results: Modelling

Our results also allow us to analyse the variation in age effect across countries for bacteria-antibiotic combinations, and we see little country-level patterns (Figure 5). As in, there is no trend for similar age-associations across bacteria-antibiotic within a country. The impact of age across different countries is correlated for some bacteria-antibiotic combinations, such as some *S. aureus* and *E. coli* resistances (Figure 6). We also look at the magnitude of the age effect across sub-regions and bacteria-antibiotic, by ranking each country by the magnitude of effect for each bacteria-antibiotic over ages 1-100, and summing across bacteria-antibiotic combinations. Investigating these by subregion show stark differences, with a higher magnitude effect of age in Southern / Eastern Europe than in Northern / Western Europe (Figure 7).

**Figure 5:**
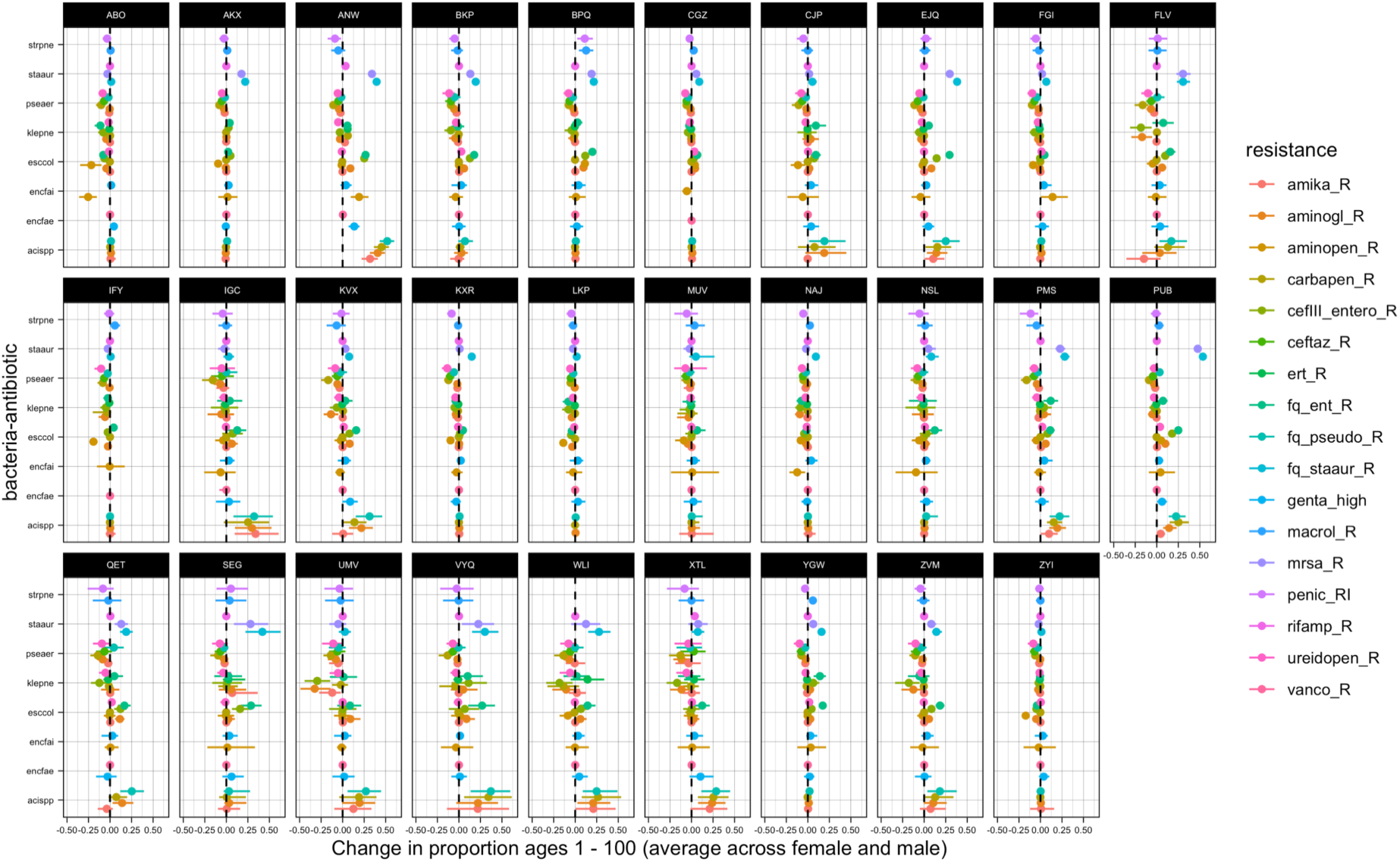
Country level age effect (difference in proportion resistant between ages 1 and 100 with 95% quantiles) by bacteria-antibiotic combination. Country labels are random anonymised three letter code used for this analysis only.

**Figure 6:**
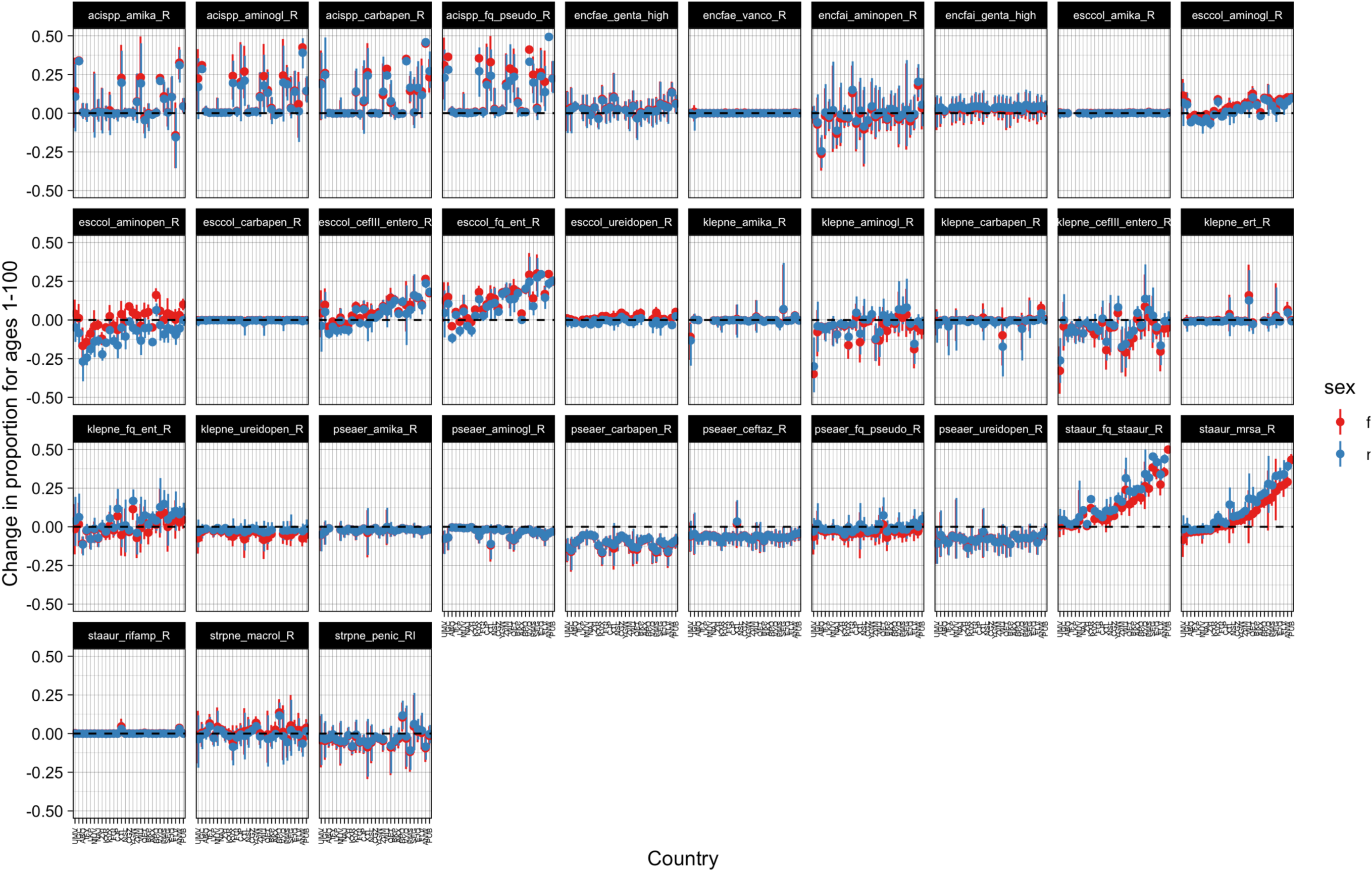
Changes in proportion for ages 1-100 by country (x axis) and sex (colour). Country labels are random anonymised three letter code used for this analysis only.

**Figure 7:**
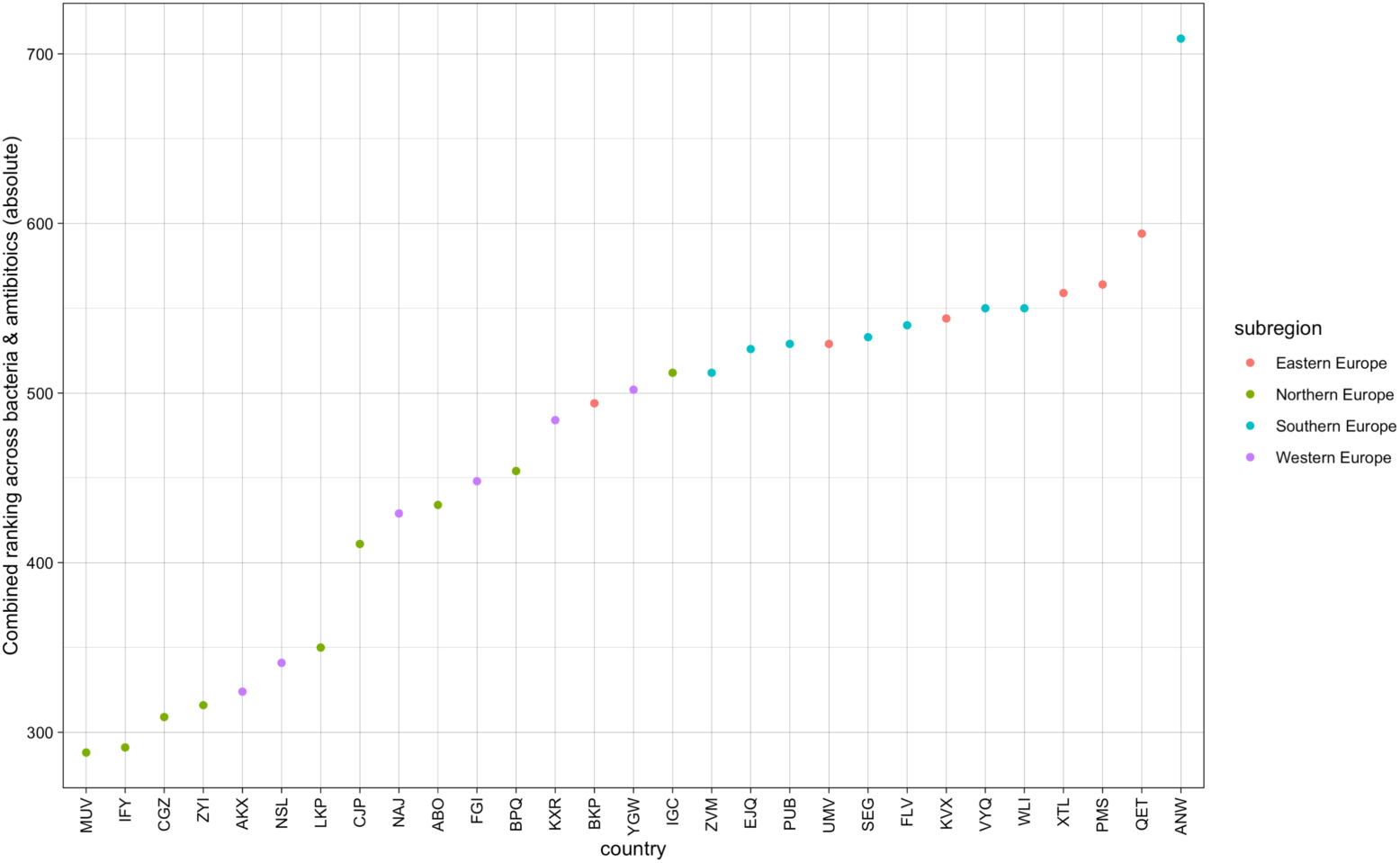
Combined ranking across bacteria-antibiotics for the magnitude of the age effects between ages 1 and 100. Country labels are random anonymised three letter code used for this analysis only.

### 6. Sensitivity analysis: data disaggregation by inpatient vs outpatient

#### Background

The original dataset includes a category of “Patient type” which recorded whether the patient at the moment the sample was taken was admitted in a hospital (“INPAT”, inpatient), or not (“OUTPAT”, outpatient). Patients that go to the hospital for dialysis or other types of day hospital care are classified as other (“o” in the data) or “unknown”.

This category could be used as a proxy for the important definition of community-vs hospital-acquired infection as the data is on the first blood and/or cerebrospinal fluid isolate from a patient. However, the standard definition splits the above classification based on the first 48hrs of patient care. Hence, some of the “inpatient” samples here will be officially “community acquired” (taken when the patient has officially been admitted but before they have been in hospital < 48hrs) (8). We therefore cannot directly correlate this patient type with hospital vs community. Moreover, not all countries report this distinction – the UK has “unknown” for all isolates.

Exploring the data pooled across all countries with data, reveals the same age-dependent incidence curve for all bacteria across patient type where substantial data exists i.e. for inpatient and outpatient classifications and not “other” (Figure 8). Incidence is lower in outpatients as would be expected. Similar patterns can be seen for individual countries and across sexes as seen without the patient type split. However, this disaggregation also showed certain countries had substantially more isolates from outpatients. This likely reflects differences in healthcare processes and data coding.

**Figure 8:**
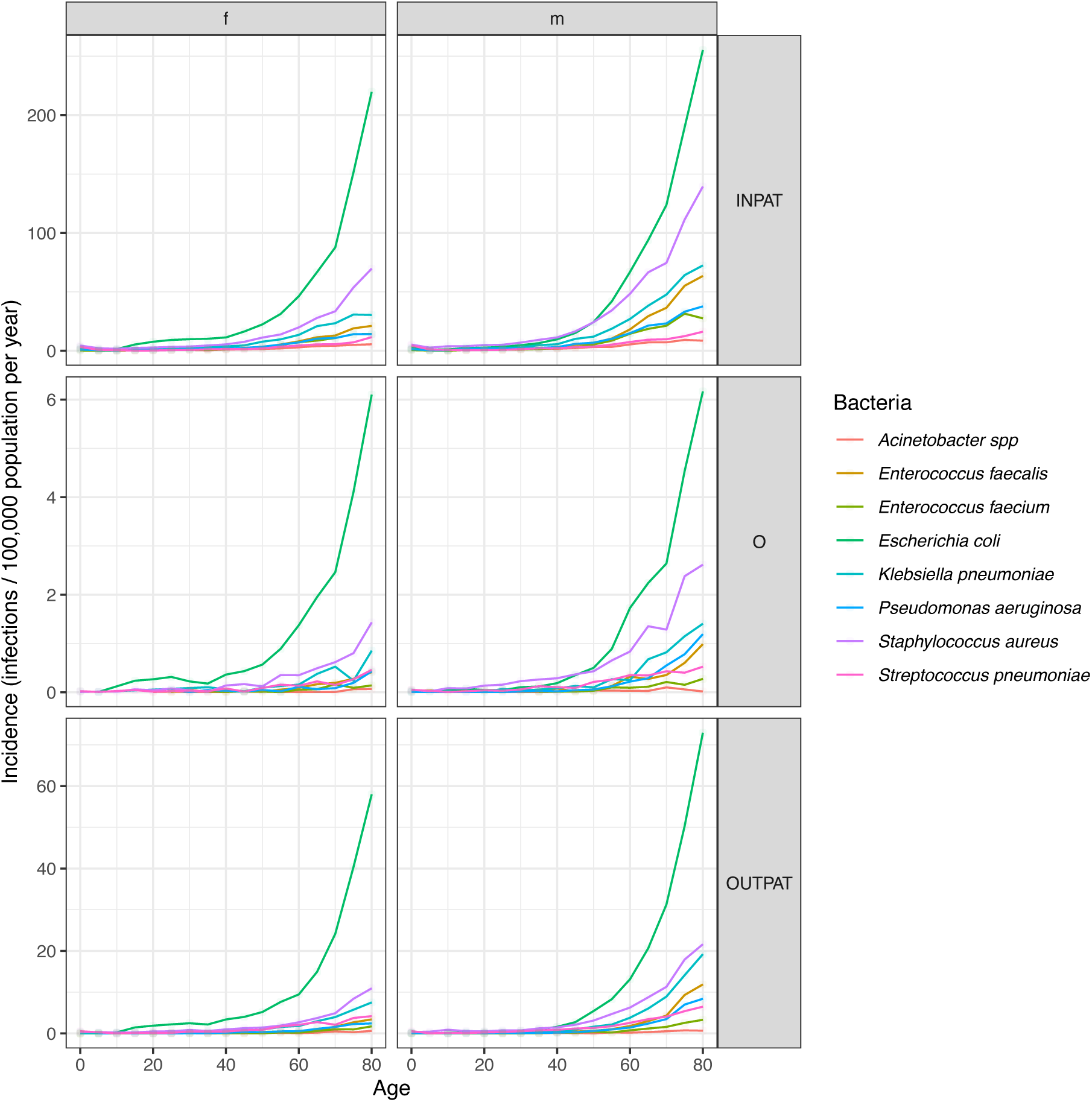
Impact of patient type (inpatient = INPAT, outpatient = OUTPAT or Other) on incidence of bloodstream infection for different bacterial types (colour) for 2019 for females (left) and males (right).

### Conclusions

The similarity of the high-income healthcare settings across Europe suggest that the above variations may be due to country level differences (“cultural factors”) in sampling and reporting rather than actual differences in bloodstream infection incidence by patient type. Hence, we have not explored resistance prevalence further by these patient types without further information on sample collection by country.

### 7. Sensitivity analysis: data disaggregation by hospital unit type

#### Background

The dataset included a category of “Hospital unit type” which recorded the hospital unit within which the patient was when the sample was taken.

The Hospital Unit Types in data are:

ED = Emergency Department, ICU = Intensive Care Unit, INFECT = Infectious Disease Ward, INTMED = Internal Medicine, O = Other, OBGYN = Obstetrics/Gynecology, ONCOL = Haematology/Oncology, PEDS = Pediatrics/neonatal, PEDSICU = Pediatrics/neonatal ICU, PHC = Primary Health Care, SURG = Surgery, UNK = Unknown, URO = Urology Ward

#### Data description

Most were internal medicine (“INTMED”, 27%) or “Unknown” (21%), followed by Emergency Department (“ED”, 14%), the “ICU” (11%) and Surgery (“SURG”, 8%).

The distribution varied substantially by country – with one country have up to 53% of isolates reported as unknown, whilst another had the up to 45% as “Internal medicine” .

The age / sex distribution of incidence results across hospital unit types and further over countries are basically the same as across all data – there is an age association, with *E. coli* being the dominant bacteria (Figure 9). There seem some issues with the paediatric data (PEDS / PEDSICU) data with some isolates having older age associated with them. The data for oncology (“ONCOL”) suggests there may be some survival bias effects (Figure 9). *S. aureus* has the second highest infection incidence across all hospital unit types except for urology (“URO”) where other bacteria have relatively equal infection incidence levels which makes sense. The data from obstetrics and gynaecology (“OBGYN”) emphasises the importance of childbirth as a risk factor for women, though with relatively low rates.

**Figure 9:**
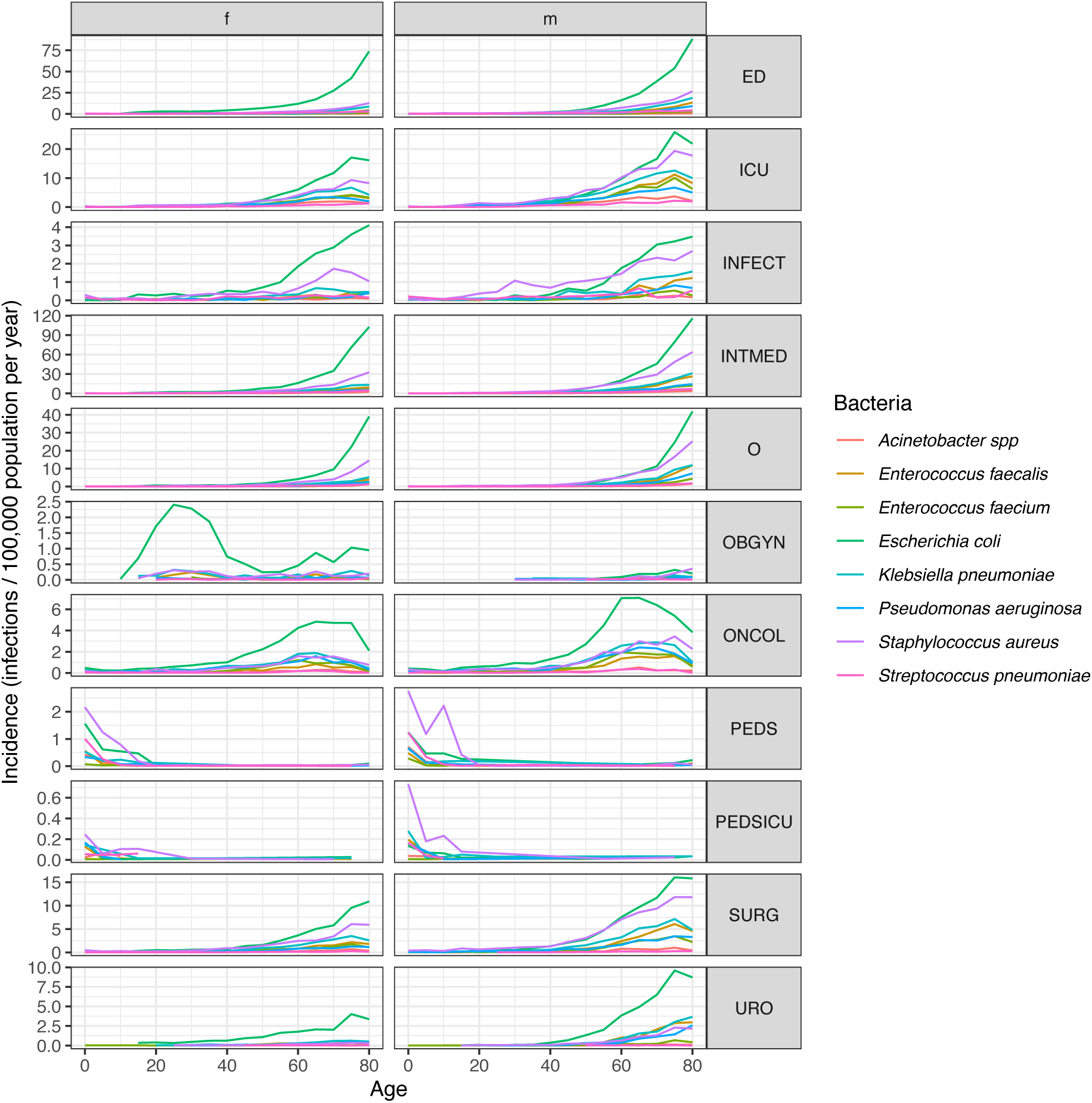
Total incidence of infection by age (x axis) and sex (columns) over all countries for different hospital unit types (rows) and bacteria (colours) in 2019. ED = Emergency Department, ICU = Intensive Care Unit, INFECT = Infectious Disease Ward, INTMED = Internal Medicine, O = Other, OBGYN = Obstetrics/Gynecology, ONCOL = Haematology/Oncology, PEDS = Pediatrics/neonatal, PEDSICU = Pediatrics/neonatal ICU, PHC = Primary Health Care, SURG = Surgery, UNK = Unknown, URO = Urology Ward

The distribution of infection incidence emphases the above patterns showing for women aged 15-40 approximately 20% of the recorded bloodstream infections are linked to stays in the obstetrics and gynaecology departments (Figure 10). In men of this age, there is a larger contribution of infections in the ICU.

**Figure 10:**
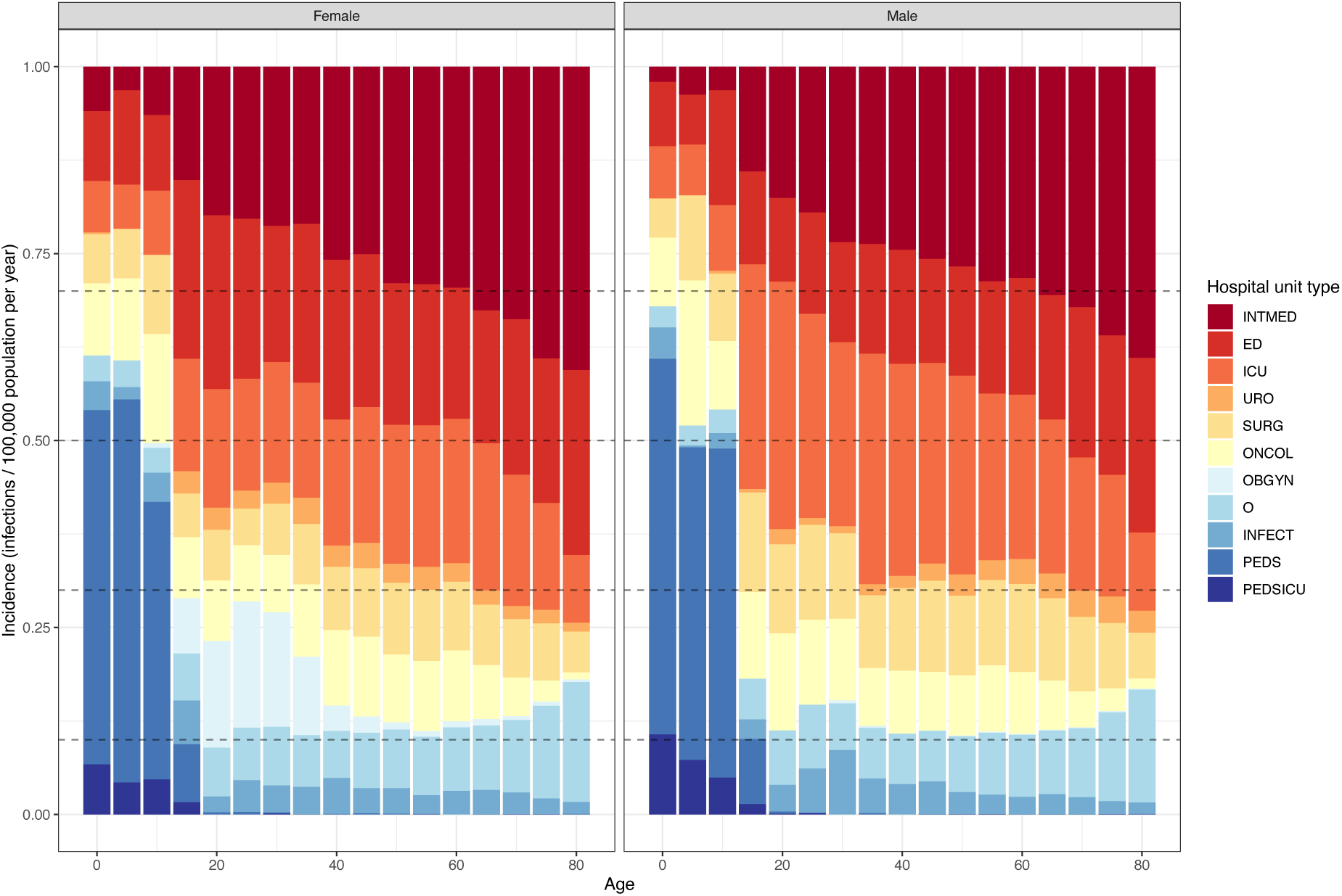
Distribution of infection incidence by hospital unit type (colour) across age (x axis) and sex (panels across Europe). ED = Emergency Department, ICU = Intensive Care Unit, INFECT = Infectious Disease Ward, INTMED = Internal Medicine, O = Other, OBGYN = Obstetrics/Gynecology, ONCOL = Haematology/Oncology, PEDS = Pediatrics/neonatal, PEDSICU = Pediatrics/neonatal ICU, PHC = Primary Health Care, SURG = Surgery, UNK = Unknown, URO = Urology Ward

#### Conclusions

The similarity of the high-income healthcare settings across Europe suggest that the above variations may be due to country level differences (“cultural factors”) in sampling and reporting rather than actual differences in bloodstream infection incidence by hospital unit type. Hence, we have not explored resistance prevalence further by these hospital unit types without further information on sample collection by country.

### 8. Sensitivity analysis: incidence

Using the minimum (instead of the actual reported or estimated coverage in 2019) estimates of the population coverage across the 2015-2019 data and estimates did not affect the sex and age patterns (Figure 11). The number of infections did increase substantially, as would be expected as coverage has been increasing for many countries, and also infections from countries that did not have coverage estimates for 2019 would be included if they had estimates for other years. On average, across all countries and years, the incidence was increased by ∼38%, with similar increases for both sexes.

**Figure 11:**
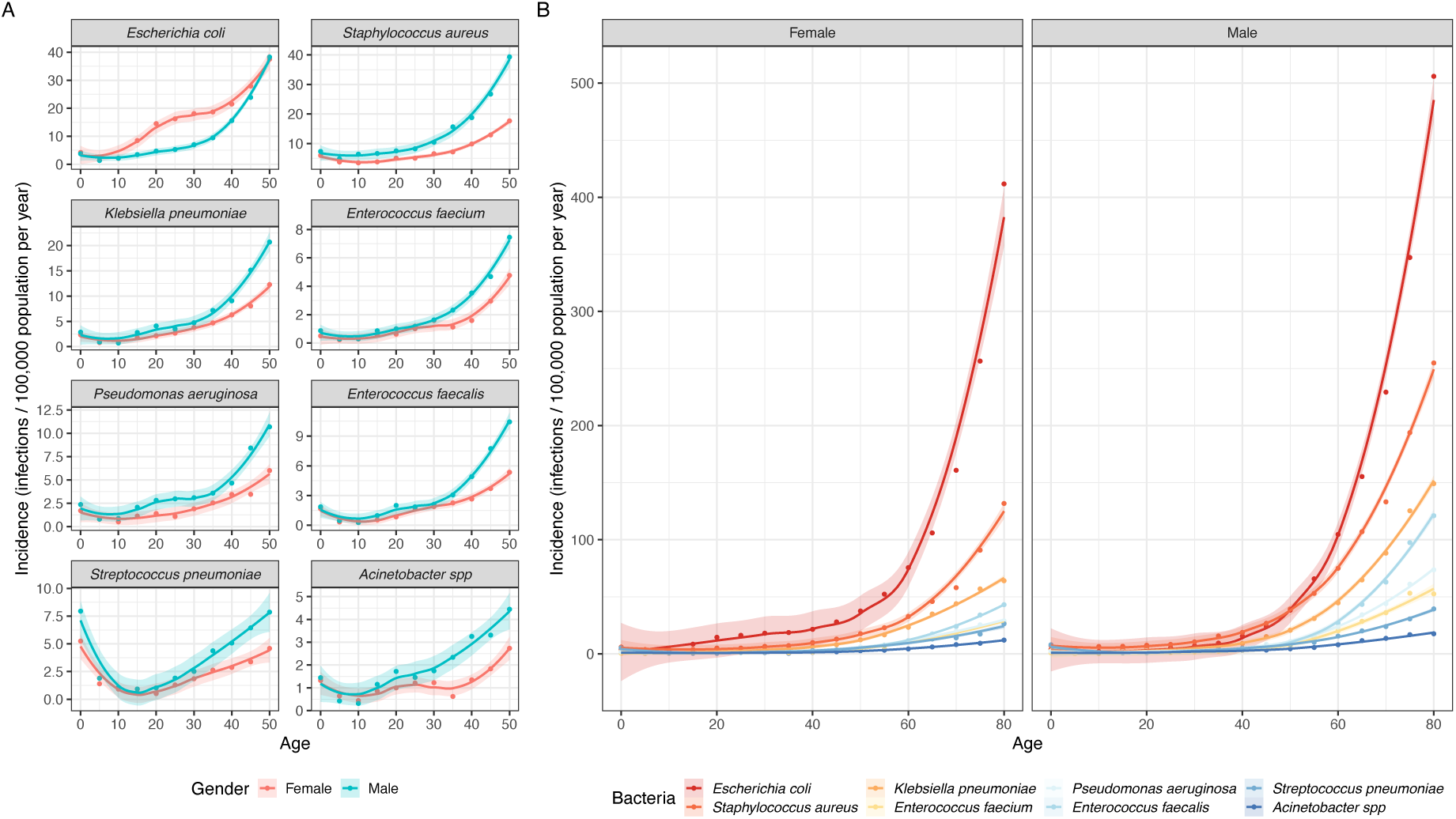
Incidence of bloodstream infections calculated using the minimum reported coverage estimates for surveillance per 100,000 population in 2019 across European countries for 8 bacterial pathogens split by (A) sex and bacteria for the first 50yrs of life and (B) sex (panel) and bacteria (colour) lifelong. Shaded areas are 95% confidence intervals using a LOESS fit. Infections in individuals younger than 0 are excluded, and those aged 80 and older are pooled into the 80yr data point. of bloodstream infections per 100,000 population per year across European countries for 8 bacterial pathogens (panel) for multiple years (colour) and split by sex (solid line = female, dashed line = male). Shaded areas are 95% confidence intervals using a LOESS fit.

### 9. Sensitivity Analysis: Priors

In our main paper models we had flat priors on all fixed effect variable parameters. In order to test the strength of our conclusions, we reran the MRSA model including regularising priors of *normal*(*0,1*) for all the fixed effect variables. We chose MRSA to run our sensitivity analysis, as this was one of the case studies in the main manuscript. Results showed no effect of the regularising priors on the estimated posterior parameters (Figure 12).

**Figure 12:**
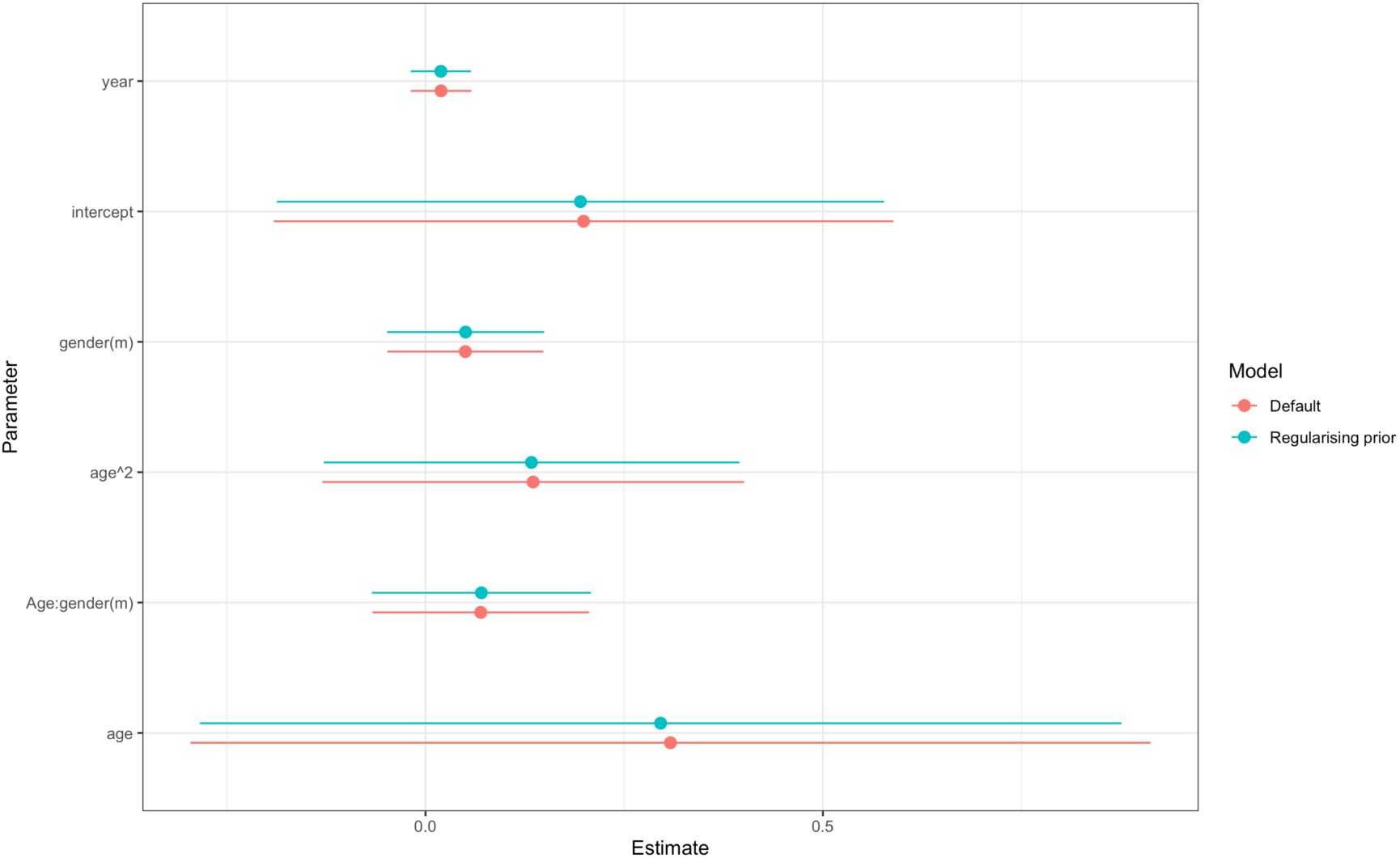
Parameter comparisons for sensitivity analysis including regularising priors. Points indicate the mean of the posterior estimate, with lines indicating the 95%CI.

### 10. Sensitivity Analysis: Aged 0

In our main analysis we excluded infants aged 0 from the data, due to large differences in their immune system and infection mechanisms. To test the impact of this, we reran the model for MRSA including individuals aged 0. We found no difference in the posterior parameter estimates for the fixed effect model variables (Figure 13A). Moreover, no substantial differences were seen in the random country level effects of age (Figure 13B).

**Figure 13:**
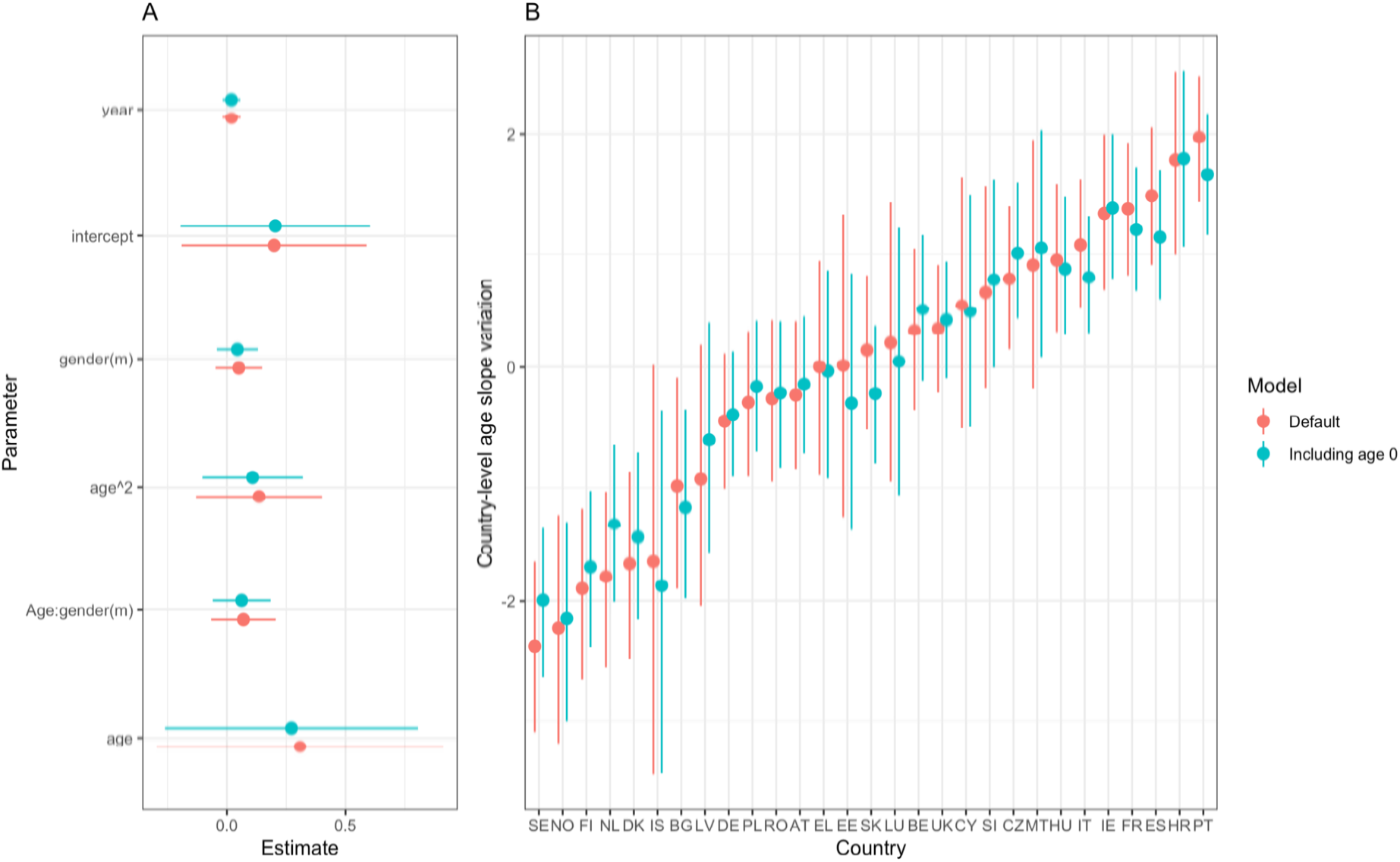
Parameter comparisons for sensitivity analysis including data from infants aged 0. Points indicate the mean of the posterior estimate, with lines indicating the 95%CI.

### 11. Sensitivity Analysis: Model philosophy

Whilst our main model used a Bayesian approach, where model variables that had no impact would have a parameter variable centred around 0, here we present a model comparison approach, for an example bacteria-antibiotic combination. In this approach, our base model (model A) did not account for sex or age impacts on resistance, but included a covariate for year, as well as random intercept effects for country and laboratory code (Equations 1:2). Our age model (model B) included additional covariates age, age-squared and a random covariate effect of age by country (Equations 3:4). This reflects the potential for age trends to be different across country due to differing policy and prescription patterns. Model C was the same as the base model, but additionally included a sex covariate (Equations 5:6). Model D was the complete model, including both age and sex terms, as in the main paper. Differences between the base model and subsequent models are shown in bold.

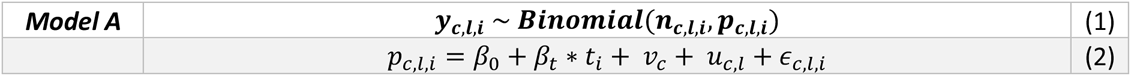

Where *y* is the resistance variable, taking a value of 0 or 1, *n* is the number of samples and *p* the probability of the sample being found to be resistant. The subscripts *c*, *l* and *i* denote country, laboratory code and grouping level. Recall that for each bacteria-antibiotic combination, our data consisted of multiple groupings of individual samples of a bacterium tested for resistance to that antibiotic. Each grouping *i* had a unique combination of country (c), laboratory code (l), sex, age and year of sample and hence a linked number of samples (*n*) and proportion resistant (*p*). *β*_0_ is the overall intercept, *β_t_* is slope coefficient for time and *t* is year. *ε_c,l,t_* is the residual error, *u_c,l_* is the level-2 random error on laboratory code and *v_c_* is the level-3 random error on country.

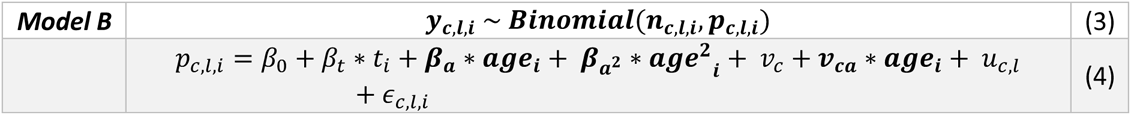

Where *β_a_* is the age effect coefficient, *β_a^2^_* is the age squared effect coefficient and **v_ca_** is the country level age effect coefficient.

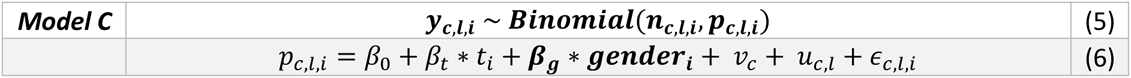

Where *β_g_* is the sex effect coefficient.

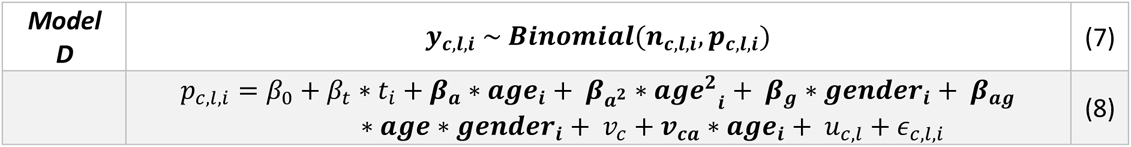

Where *β_ag_* is the sex and age interaction coefficient.

We ran this set of models for MRSA (coefficient estimates in Figure 14A). Leave-one-out cross validation (approximated by the Pareto smoothed importance-sampling method) (LOOIC) and the widely applicable information criterion (WAIC) were used to compare the predictive ability of the models.

**Figure 14:**
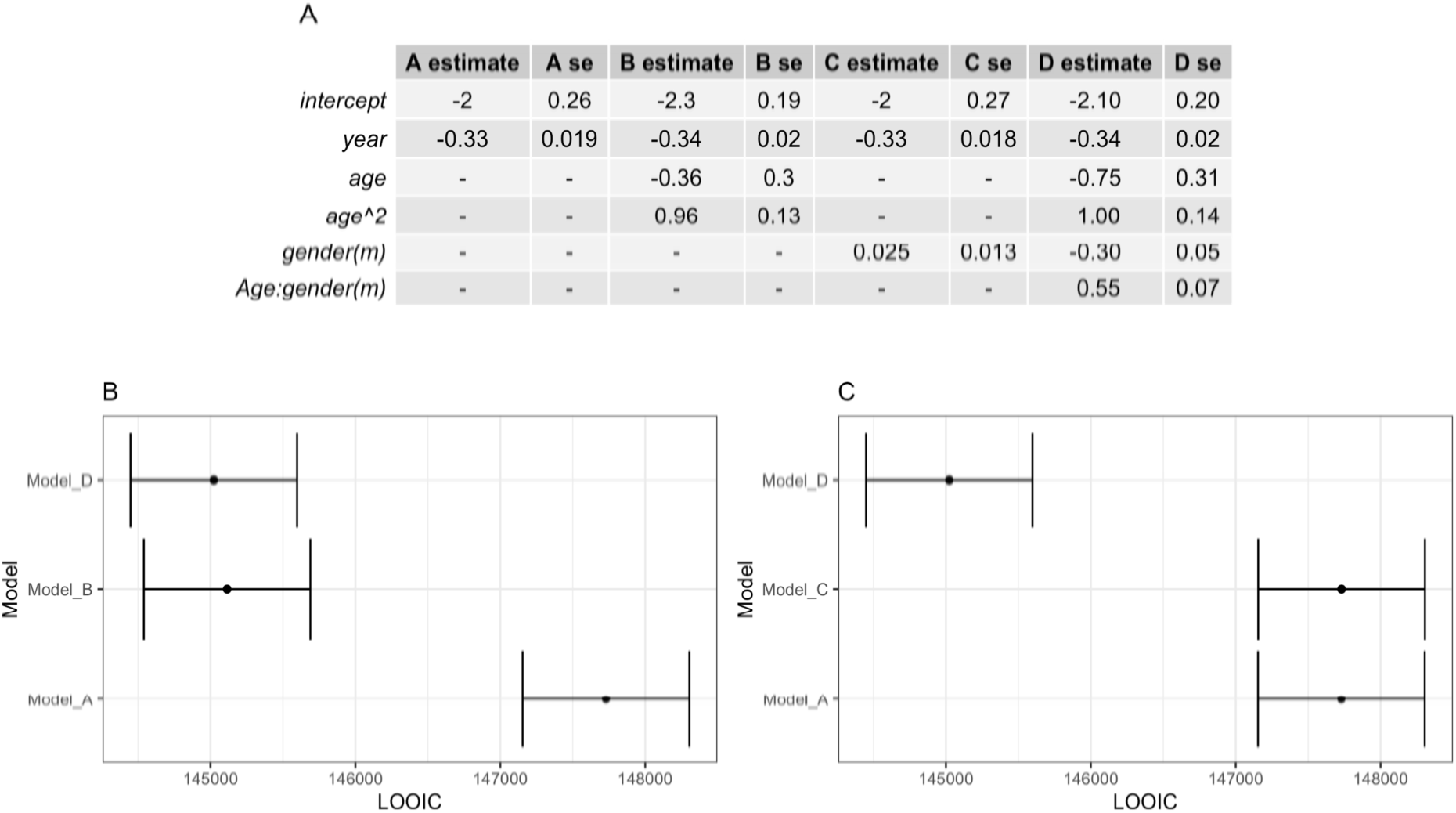
Philosophical sensitivity analysis. A) Comparison of posterior parameter values across models. B) LOOIC for nested models with age. C) LOOIC for nested models with gender.

Due to model nesting, we compared A-B-D and A-C-D separately. Including age in the base model A had a large impact on the LOOIC (Figure 14B, Model B vs Model A), and adding sex on top of that resulted an additional small improvement (Model D vs Model B). However, adding sex to the base model did not improve the LOOIC (Figure 14C, Model C vs Model A). Adding in age as well as sex had significant improvements on the LOOIC (Model D vs Model C). These results indicate that Model D, the one used in our main analysis, would be chosen as the best model to use in the model comparison approach for MRSA.

